# Machine learning for the prediction of spontaneous preterm birth using early second and third trimester maternal blood gene expression: A Cautionary Tale

**DOI:** 10.1101/2024.09.11.24313497

**Authors:** Kylie K Hornaday, Ty Werbicki, Suzanne C Tough, Stephen L Wood, David W Anderson, Constance H Li, Donna M Slater

**Affiliations:** Department of Physiology and Pharmacology, Cumming School of Medicine, University of Calgary, Calgary, Alberta, Canada; Department of Community of Health Sciences, Cumming School of Medicine, University of Calgary, Calgary, Alberta, Canada; Department of Pediatrics, Cumming School of Medicine, University of Calgary, Calgary, Alberta, Canada; Department of Obstetrics and Gynaecology, Cumming School of Medicine, University of Calgary, Calgary, Alberta, Canada; Department of Science, Langara College, Vancouver, British Columbia, Canada; Department of Biochemistry and Molecular Biology, Cumming School of Medicine, University of Calgary, Alberta, Canada

## Abstract

Spontaneous preterm birth (sPTB) remains a significant global health challenge and a leading cause of neonatal mortality and morbidity. Despite advancements in neonatal care, the prediction of sPTB remains elusive, in part due to complex etiologies and heterogeneous patient populations. This study aimed to validate and extend information on gene expression biomarkers previously described for predicting sPTB using maternal whole blood from the All Our Families pregnancy cohort study based in Calgary, Canada. The results of this study are two-fold: first, using additional replicates of maternal blood samples from the All Our Families cohort, we were unable to repeat the findings of a 2016 study which identified top maternal gene expression predictors for sPTB. Second, we conducted a secondary analysis of the original gene expression dataset from the 2016 study using five modelling approaches (random forest, elastic net regression, unregularized logistic regression, L2-regularized logistic regression, and multilayer perceptron neural network) followed by external validation using a pregnancy cohort based in Detroit, USA. The top performing model (random forest classification) suggested promising performance (area under the receiver operating curve, AUROC 0.99 in the training set), but performance was significantly degraded on the test set (AUROC 0.54) and further degraded in external validation (AUROC 0.50), suggesting poor generalizability, likely due to overfitting exacerbated by a low feature-to-noise ratio. Similar performance was observed in the other four learning models. Prediction was not improved when using higher complexity machine learning (e.g. neural network) approaches over traditional statistical learning (e.g. logistic regression). These findings underscore the challenges in translating biomarker discovery into clinically useful predictive models for sPTB. This study highlights the critical need for rigorous methodological safeguards and external validation in biomarker research. It also emphasizes the impact of data noise and overfitting on model performance, particularly in high-dimensional omics datasets. Future research should prioritize robust validation strategies and explore mechanistic insights to improve our understanding and prediction of sPTB.

## Introduction

Preterm birth, defined as delivery of a live infant prior to 37 weeks of gestation, occurred in approximately 13.4 million births worldwide in 2020, and is a significant contributor to mortality and morbidity in neonates and children under five [1, 2]. Child mortality related to PTB complications has declined since 2000, in part due to advancements in treatments for neonatal complications of prematurity such as respiratory distress syndrome. However, an estimated 900,000 PTB-associated deaths of children under five still occurred in 2019 worldwide [3].

Approximately one third of preterm births occur because of known maternal or fetal indications, while the remaining two thirds occur following spontaneous onset of labour and/or premature rupture of the fetal membranes (collectively termed spontaneous or sPTB). Most sPTB are without known indication, making prediction and subsequent clinical management of risk, challenging [2]. As one of the great obstetrical syndromes, considerable efforts have aimed to identify predictive biomarkers of sPTB, however none so far have emerged to have clinical utility, possibly due to heterogeneity within both patient populations and preterm birth phenotypes, as well as risk of bias within study design [4–7]. Methodological safeguarding and appropriate validation of models is important to determine feasibility, repeatability, robustness, and generalizability of prediction [8–11]. Best practices for prediction modelling are well defined in the literature [12, 13], and primary research articles reporting external validation of prediction models have been increasingly published over the last five years [14–17]. However, in the reproductive field, studies externally validating prediction models are limited [10, 18, 19]. Thus, we sought herein to repeat and validate previous findings on prediction of sPTB, which identified a predictive relationship between gene expression biomarkers and sPTB.

Gene expression biomarkers have been identified in maternal whole blood for the prediction of sPTB, which presents a promising avenue for minimally invasive prediction as peripheral blood can reflect global and uterine physiological and immunological changes during pregnancy [20]. One example includes eight genes, *LOC100128908, MIR3691, LOC101927441, CST13P, ACAP2, ZNF324, SH3PXD2B, TBX21* that were identified as significantly predictive of sPTB (65% sensitivity and 88% specificity after adjusting for history of abortion and anaemia) in a stepwise logistic regression model published in 2016 by Heng *et al.* [21]. These gene expression biomarkers were originally identified using an Affymetrix chip microarray analysis of maternal whole blood from the All Our Families pregnancy cohort based in Calgary, Canada [21]. The All Our Families pregnancy cohort presents a rare opportunity for testing experimental repeatability, as maternal blood samples were collected and stored in four separate PAXgene RNA tubes, two were used for the original study (22), and one for validating RNA quality and integrity [22], leaving a remaining fourth sample for experimental validation. The study herein sought to use the additional PAXgene tube to repeat and validate this predictive model to test feasibility for clinical use.

It is also important to note that prediction algorithms that match too closely to the training data, in other words, suffer from overfitting, are not generalizable to other populations, which is one of the major limitations of prediction modelling. This problem is exacerbated by small or non-representative training sets, where patterns identified may not be meaningfully associated with the outcome, or constitute “noise”, and thus the prediction does not translate effectively beyond the original training observations. This stresses the importance of external validation in order to identify robust, generalizable models to meaningfully push forward the prediction of preterm birth. Therefore, an external pregnancy cohort based in Detroit, USA, which collected maternal blood gene expression data from comparable timepoints in pregnancy, was identified, presenting a unique opportunity for external validation to determine the generalizability of prediction [23].

The original 2016 study used a logistic regression-based model; however, we hypothesized that machine learning approaches could improve predictive performance. Machine learning and other complex data analysis methods are particularly well suited for mining high dimensional datasets, such as transcriptomic datasets, as they do not require the data to adhere to any *a priori* assumptions [24]. Machine learning allows for the identification of non-obvious, interactive, complex, and/or non-linear patterns which can go undetected when using traditional statistical linear models [24, 25]. These patterns can be leveraged, both toward outcome prediction (something highly valuable for complex medical conditions such as preterm birth), and characterizing underlying disease mechanisms [26, 27]. This is particularly enticing, as the underlying causes of sPTB remain poorly understood.

The overarching aim is to explore the repeatability, generalization, and robustness of a prediction model for sPTB using maternal blood gene expression biomarkers, with emphasis on external validation of prediction models for sPTB. Additionally, given the high complexity of preterm birth, we wanted to explore whether additional learning algorithms could improve prediction over traditional statistical learning. Specific aims are as follows:

1. To test the predictive utility of the gene expression biomarkers, *LOC100128908, MIR3691, LOC101927441, CST13P, ACAP2, ZNF324, SH3PXD2B, TBX21* previously identified in maternal blood as biomarkers of spontaneous preterm birth [21].
2. To identify a predictive biomarker(s) for spontaneous preterm birth using maternal blood gene expression data using machine learning best practices and multiple learning approaches.

## Methods

### Biological samples and validation of top biomarkers

To test the reproducibility of top biomarkers identified in the literature, historical biological samples were collected from the All Our Families cohort [28–30]. In brief, participants were recruited between May 1^st^, 2008, and December 31^st^, 2011, at <25 weeks gestation and provided consent for blood sample collection, and completed questionnaires including information related to demographics, emotional and physical health. Participants provided informed written consent at the time of recruitment from healthcare offices, community and through Calgary Laboratory services, and were provided copies of their consent forms for their records. This study was approved by the Conjoint Health Research Ethics Board at the University of Calgary #REB15-0248 Predicting Preterm Birth Study. Biological samples were collected at two points in pregnancy, timepoint 1 (T1) at 17-23 weeks gestation and timepoint 2 (T2) 28-32 weeks gestation. Maternal whole blood was collected directly into four separate PAXgene blood RNA tubes which were then stored at -80°C prior to RNA isolation (PAXgene Blood RNA Kit, Qiagen). Samples were de-identified prior to data collection. Two tubes were previously used for the original prediction modelling and biomarker identification by Heng et al., [21], a third tube was used to assess RNA integrity in long term storage [22], and a fourth was collected from storage from August 16^th^ to November 3^rd^, 2018 for use in the current study. For the current study, n=47 participants who subsequently had a sPTB (<37 weeks) were included (n=44 T1, n=42 T2 samples), in addition to n=45 participants who had a healthy term (38-42 weeks) delivery (n=40 T1, n=44 T2). A total of n=13 samples were missing from storage or insufficient sample remaining (n=3 T1 sPTB, n=3 T2 sPTB, n=5 T1 term, n=2 T2 term), and n=2 T2 samples in the sPTB group were not included as delivery occurred prior to the second sample collection. Maternal blood samples were collected, and RNA was isolated according to manufacturer’s instructions (RNAeasy minikit, Qiagen). The following genes were measured using a probe-based assay (Quantigene, Invitrogen, ThermoFisher Scientific), which uses identical probes to an Affymetrix microarray chip: *LOC100128908* (*LMLN2), LOC101927441, CST13P, ACAP2, ZNF324, SH3PXD2B, TBX21.* Due to limitations with measuring microRNA (miRNA) using the assay, *MIR3691* was not measured. The same genes were also validated using bulk RNA sequencing (Illumina mRNAseq following poly-A capture, NovaSeq S4 200 cycle). Sequences were aligned to the human reference genome GrCH38 using STAR to produce reads per gene tables [31].

### Data availability

The accession number for the gene expression data from the Calgary cohort and Detroit cohort described herein are Gene Expression Omnibus GSE59491 and GSE 1494490, respectively. Scripts for data preprocessing, differential expression analysis, feature selection, and model training are available at https://github.com/tywerbicki/Slater_Lab_SPTB.

### Population and expression dataset

To test whether additional learning models could improve predictive performance, a secondary analysis of the previously published maternal blood microarray data [21], was conducted. Gene expression data was downloaded as log2 robust multi-array average (RMA) values from the National Center for Biotechnology Information Gene Expression Omnibus (accession number: GSE59491) (All Our Families, AOF- Calgary cohort) [21]. The RMA preprocessing, including background correction and normalization was previously conducted [21, 23]. The dataset used herein contains high-throughput expression data from n=165 subjects (n=51 sPTB, n=114 matched term delivery controls) nested within the Calgary AOF cohort.

Matched gene expression data from two timepoints, 17-23 weeks (T1) and 28-33 weeks (T2) was available for each participant. Two observations from the sPTB group were removed from the dataset as these deliveries occurred prior to the T2 collection and therefore have only one expression dataset.

An external dataset was additionally identified from a pregnancy cohort based in Detroit, USA [23], which collected maternal blood samples at comparable timepoints for gene expression analysis, (accession number: GSE149440), and was used for external validation of the model.

Participants that had at least two matched blood samples collected within the same two timeframes (T1 and T2) were selected from within the Detroit cohort dataset for analysis, for a total of n=98 subjects (n=34 sPTB and n=64 matched term delivery controls) included for external validation.

The Calgary dataset was randomly split into ten cross validation folds: each tenth acted as a as a test set for the preprocessing, differential expression analysis, feature selection, and model training done on the remaining 90% of the data. While there is no true hold out set for the Calgary cohort, models are externally validated on the Detroit dataset. A schematic outlining the analytical pipeline is provided in Figure 1.

**Figure 1.**
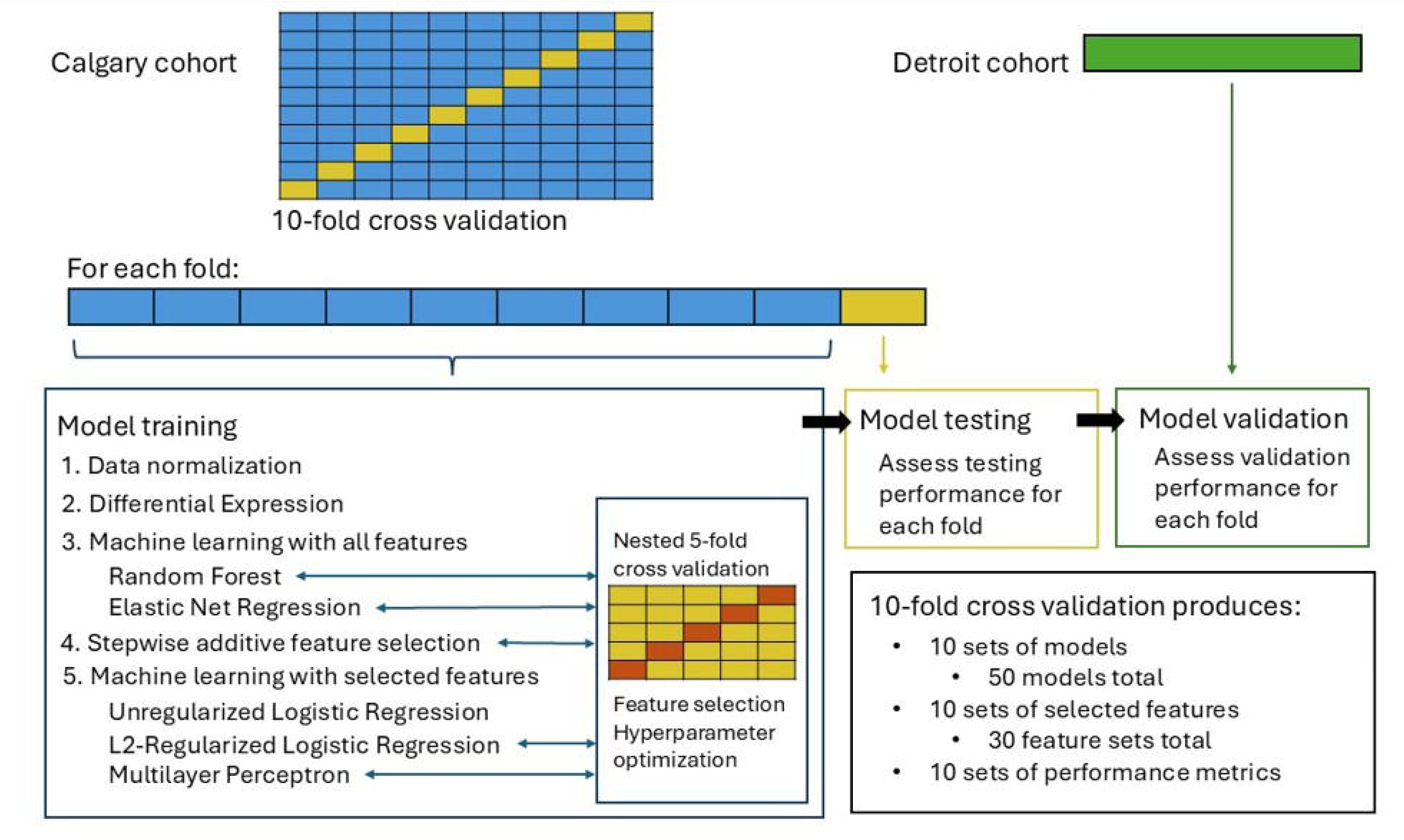
Schematic of analytical pipeline.

### Differential expression analysis

Differential expression was performed on the Calgary dataset with 10-fold cross validation. Each training fold contains 85 term and 35 sPTB labeled patients, and each testing fold contains 29 term and 12 sPTB labeled patients. For each fold, features with low and excessively high variance were filtered from analysis (feature kept if standard deviation between [0.001, 3]).

Features were also removed if microarray expression was lower than a value of 5 in more than 35 samples. Because T1 and T2 are modelled together, only genes with measurements at both time points were kept after filtering.

For each remaining gene, microarray expression was modelled against a variable combining timepoint with birth outcome using a linear model (lmFit). The correlations of repeated measures (duplicateCorrelation) were estimated and specified in the linear model. Further, an empirical Bayes model was used to adjust the linear model estimates for more robust estimates.

Differential expression results were explored using the following comparisons:

- sPTB group compared to term group at T1
- sPTB group compared to term group at T2
- T1 compared to T2 in sPTB group
- T1 compared to T2 in term group
- dT (T2-T1) in sPTB compared to term

The Benjamini-Hochberg procedure was used to perform FDR correction. Genes with a family-wise error rate less than 0.05 for any of the above comparisons were considered differentially expressed and kept for downstream modelling.

### Feature selection analysis

Feature selection was performed using a stepwise additive selective (SAS) approach with a logistic regression model. A cross-validation area under the receiver operator curve (AUROC) was used as the metric for evaluating model improvement. The full set of input features includes three features describing each differentially expressed gene: the log2 intensity at T1 (T1), the log2 intensity at T2 (T2), and the difference between these measurements (T2-T1, or dT). The possibility of interactions between each of these features was included by using a polynomial model to generate pairwise feature combinations. The number of interaction terms was restricted to a maximum of five.

The algorithm used here starts feature selection with an empty base logistic regression model. At each iteration, the addition of each available feature to the current model was assessed using a nested five-fold cross-validation scheme. The validation AUROC was calculated for each cross-validation fold and averaged across the folds to evaluate the performance for adding that feature to the current model. The feature corresponding with the highest average AUROC was added if the marginal improvement to the current model met a predetermined threshold (ε) of 0.015. If this minimum improvement in AUROC was not met, the next iteration re-run with the unchanged current model. If there was no change in the current model for three iterations, feature selection terminated and returned the features in the base model. Otherwise, the current model was updated with the selected feature for the next iteration, and feature selection continued until a maximum number of 30 predictive features were selected.

### Model training and testing

The identified predictive features in each fold were used to train three models: an unregularized logistic regression (LR), an L2-regularized logistic regression (ridge regression, LR-reg), and a multilayer perceptron (MLP) neural network. The resultant three models were assessed for predictive performance by fitting them on the cross-validation test set and the external (Detroit) dataset. The Python Scikit-Learn implementation of logistic regression was used with a Newton-Conjugate gradient and a maximum of 10,000 iterations. For the L2- regularized logistic regression, a nested 5-fold cross-validation was used for hyperparameter tuning on the penalty value (regularization strength).

The PyTorch, Scikit-Learn and Ray Tune Python libraries were used for MLP implementation. The MLP has three layers: two hidden layers and one output layer. We used a Leaky Rectified Linear Unit (Leaky ReLU) activation function, dropout regularization, and performed batch normalization for each hidden layer. Nested five-fold cross-validation and Ray Tune was used to optimize the following hyperparameters: the number of units in the first and second hidden layers (l1, l2), dropout probabilities for the first and second hidden layers (p1, p2), the learning rate (lr), weight decay (L2 regularization), and the training batch size. We set Ray Tune to explore 500 hyperparameter configurations (num_samples=500) and used a reduction factor of four (η =4) for more aggressive pruning. To select the best trained model, we compared validation loss and AUROC. Training for the optimized MLP model was limited to 250 epochs.

Additionally, we fit two machine learning models after differential expression analysis but prior to formal feature selection: an elastic net logistic regression (LR-ELN) and a random forest classifier (RF). These models have internal feature selection and provide comparison feature sets for our stepwise additive feature selection approach. The elastic net model was fit using the Scikit-Learn implementation with a saga solver and a maximum of 10,000 iterations. The random forest model was fit using the Scikit-Learn implementation (RandomForestClassifier). Hyperparameters for both models (elastic net: mixing parameter, regularization strength; random forest: n trees, max depth, min samples to split, min samples per leaf, feature selection at split) were optimized using nested 5-fold cross-validation.

## Results

### Population demographics

Demographic characteristics of the population used for biomarker validation are described in Table 1. Participants with sPTB did not significantly differ from the term group in age, smoking status, alcohol use during pregnancy, history of abortion, history of PTB, gravidity or parity.

**Table 1.** Patient demographic and clinical characteristics.

|  | **sPTB**<br>**(n=47)** | **Term**<br>**(n=45)** |
| --- | --- | --- |
| **Maternal age (years)** | 31.5[30.2-32.8] | 31.8[30.6-33.0] |
| **Maternal ethnicity** |  |  |
| **White** | 37, 79[64-89]% | 35, 78[63-89]% |
| **Non-white** | 10, 21[11-36]% | 10, 22[11-37]% |
| **Smoking during pregnancy** |  |  |
| **Yes** | 5, 11[4-23]% | 10, 22[11-37]% |
| **No** | 36, 77[62-88]% | 34, 76[60-87]% |
| **Unknown** | 6, 13[5-26]% | 1, 2[0.06-12]% |
| **Alcohol during pregnancy** |  |  |
| **Yes** | 17, 36[23-52]% | 24, 53[38-68]% |
| **No** | 22, 47[32-62]% | 18, 40[26-56]% |
| **Unknown** | 8, 17[8-31]% | 3, 7[1-18]% |
| **History of abortion** |  |  |
| **Yes** | 4, 9[2-20]% | 8, 18[8-32]% |
| **No** | 43, 91[80-98]% | 37, 82[68-92]% |
| **History of PTB** |  |  |
| **Yes** | 8, 17[8-31]% | 2, 4[0.5-15]% |
| **No** | 39, 83[69-92]% | 43, 96[85-99]% |
| **Gravidity** | 1.9[1.6-2.2] | 1.9[1.7-2.2] |
| **Parity** | 0.6[0.3-0.8] | 0.5[0.4-0.7] |
| **Gestational age at delivery (weeks)** | 33.9[33.2-34.6] | 39.2[38.9-39.4] |

Values are represented as mean[95% confidence interval] for continuous variables or n, %[95% confidence interval] for categorical variables.

### Validation of biomarkers of preterm birth

Of the biomarkers measured, only five of seven were detectable in the study population using both a probe-based assay (Table 2, S1 Table) and RNA sequencing (S1 Table). As both the probe-based assay and RNA sequencing exhibited similar patterns, the following analysis was conducted on the probe-based assay data. Samples were tested at four concentrations (1.875, 3.75, 6.25, 25ng/uL RNA standards). Biomarkers *CST13P* and *LMLN2* were below the limit of detection (<LOD) in over 50% of the population (68% and 56% respectively) at all concentrations and thus were excluded from further analysis. One sample (term T2) was <LOD across all biomarkers, which was likely a technical issue with sample processing and thus excluded. Levels of *SH3PXD2B* were <LOD in 22% of the population and those <LOD were assigned as one half of the basement level (3 MFI, mean fluorescence index units). The remaining four biomarkers were present above the limit of detection in all samples.

**Table 2.**
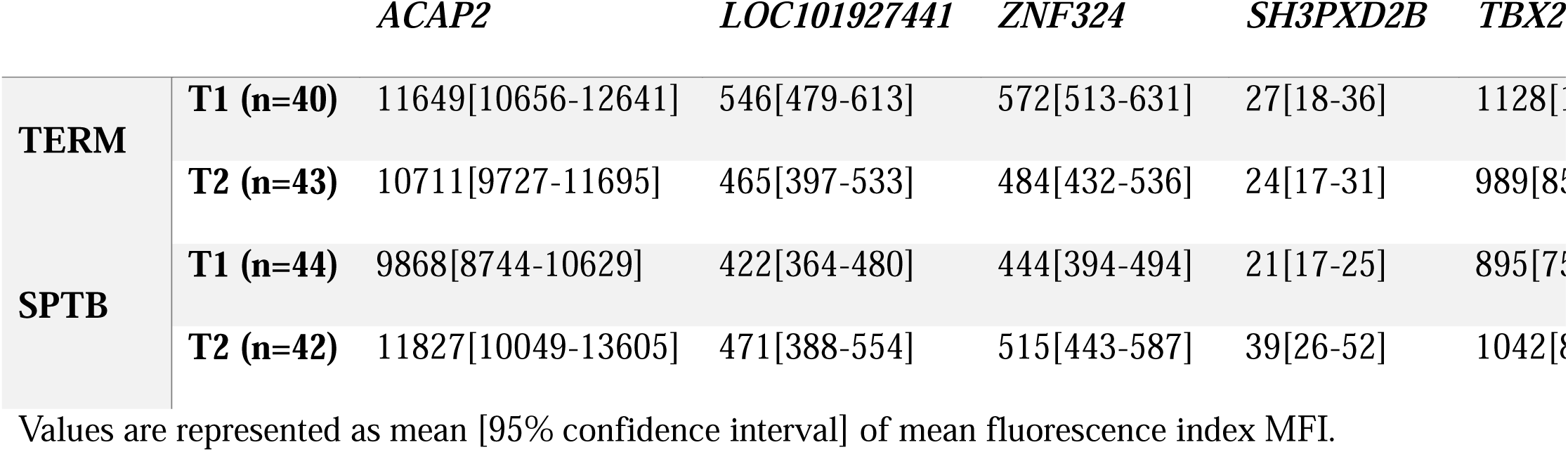
Biomarker levels in maternal blood.

Four of the five measured biomarkers, *ACAP2* (p=0.0068)*, LOC101927441* (p=0.0082), *ZNF324* (p=0.0019), and *TBX21* (p=0.0182) exhibited significantly lower levels in the sPTB group compared to the term group at T1, and not at T2. When assessing biomarkers as a measurement of T2/T1 ratios, *ACAP2* (p=0.0074)*, LOC101927441* (p=0.0273), *ZNF324* (p=0.0170), *TBX21* (p=0.0119) ratios were significantly higher in the sPTB group than the term group, suggesting a greater trajectory of increased expression through gestation in those with sPTB (Figure 2). Though we do observe some differences in biomarker levels between sPTB and term samples, only five of the eight originally identified biomarkers [21] could be measured using the same population and methodology, and only four exhibited significant differences between term and preterm groups.

**Figure 2.**
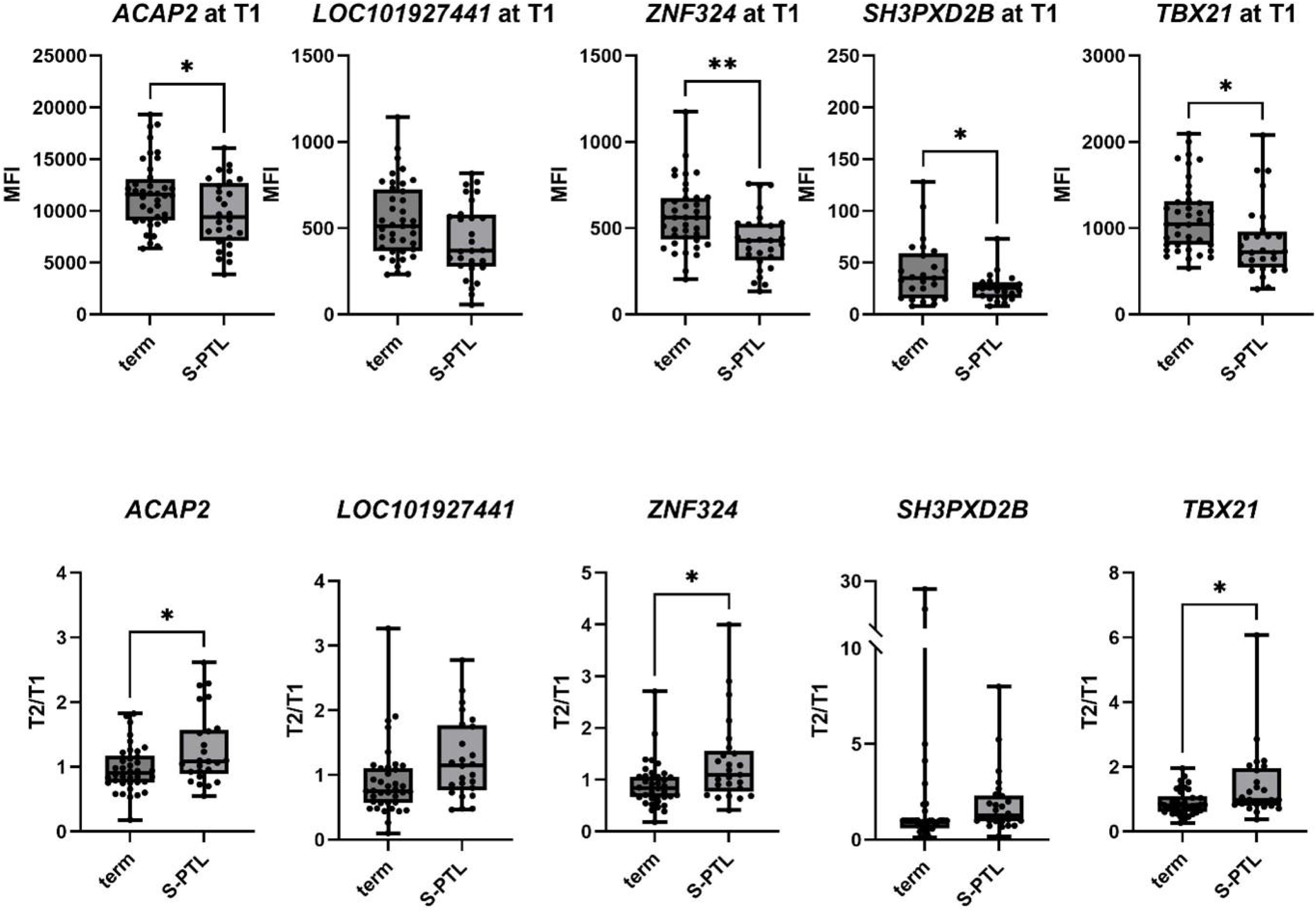
Biomarkers of preterm birth. Values are reported as mean fluorescence index (MFI) at either timepoint or a ratio of MFI values at T2 over T1. Analysed by one-way ANOVA followed by Dunnett correction for multiple comparisons. Box and whisker plots represent minimum, maximum and median values. *p-value<0.05, **p-value<0.01. *ACAP2*: ArfGAP with coiled-coil, ankyrin repeat and PH domains 2, *ZNF324*: zinc finger protein 324. *SH3PCD2B*: SH3 and PX domains 2B, *TBX21*: T-box transcription factor 21.

### Differential expression analysis

Differential expression analysis identified 1108 genes that were differentially expressed in at least one comparison, for a total of 3324 gene features (T1, T2 and T2-T1 for each gene) kept for downstream modelling (S2 Tables).

### Feature selection

The stepwise additive selection algorithm (SAS) identified 73 gene features which were selected for downstream modelling using unregularized logistic regression, an L2-regularized logistic regression, and a multilayer perceptron (MLP) neural network (S3 Table). The top (rank #1) gene features for each iteration of cross validation (two times five-fold cross validation for a total of ten iterations), are represented in Table 3. Notably, the top predictive genes did not show consistency in ranking across iterations, and no features were selected more than two times in ten iterations (range 1-2). For example, the top-most predictive feature identified by the SAS algorithm, *MLPL51,* which encodes a mitochondrial ribosomal protein, at T1 was ranked #1 most predictive feature in two iterations but was assigned a score of zero (uninformative) the remaining eight iterations, indicating that top features are not robust to noise in the dataset.

**Table 3.** Top features identified by the stepwise additive feature selection algorithm.

| Feature | Number of times selected | Average Rank | Gene name |
| --- | --- | --- | --- |
| *MRPL51* at T2 | 2 | 1 | mitochondrial ribosomal protein L51 |
| *ZNF136* at T2 | 2 | 1.5 | zinc finger protein 136 |
| *TRBJ2-6* at T1 | 1 | 1 | T cell receptor beta joining 2-6 |
| *DHRS7B* at T1 | 1 | 1 | dehydrogenase/reductase member 7B |
| *NSRP1* at T2 | 1 | 1 | nuclear speckle splicing regulatory preotin 1 |
| *C2CD2* at T1 | 1 | 1 | C2 calcium dependent domain containing 2 |
| *NCALD* at T1 | 1 | 1 | neurocalcin delta |
| *ZFP14* at T2 | 1 | 1 | zinc finger protein 14 |
| *NOLC1* at T1 | 1 | 1 | nucleolar and coiled-body phosphoprotein 1 |

Rows represent individual gene features. Number of times selected indicates the number of times each feature was selected by the algorithm across ten iterations (2 times five-fold cross validation). Average rank indicates the average rank assigned to each feature each time it was selected, with 1 being the highest possible rank.

The elastic net logistic regression model and random forest classifier were fit after differential expression analysis but prior to formal feature selection with SAS, as these models perform internal feature selection. We identified 45 gene features selected by the elastic net in >5 iterations out of 10 cross validation folds, only 10 of which were the same features identified by SAS. The random forest classifier selected 77 gene features in >5 out of 10 iterations, only 6 of which were the same features identified by SAS (Figure 3). Notably, only four features were identified by all three feature selection approaches (Table 4). A full list of features identified is available in S2 Tables.

**Figure 3.**
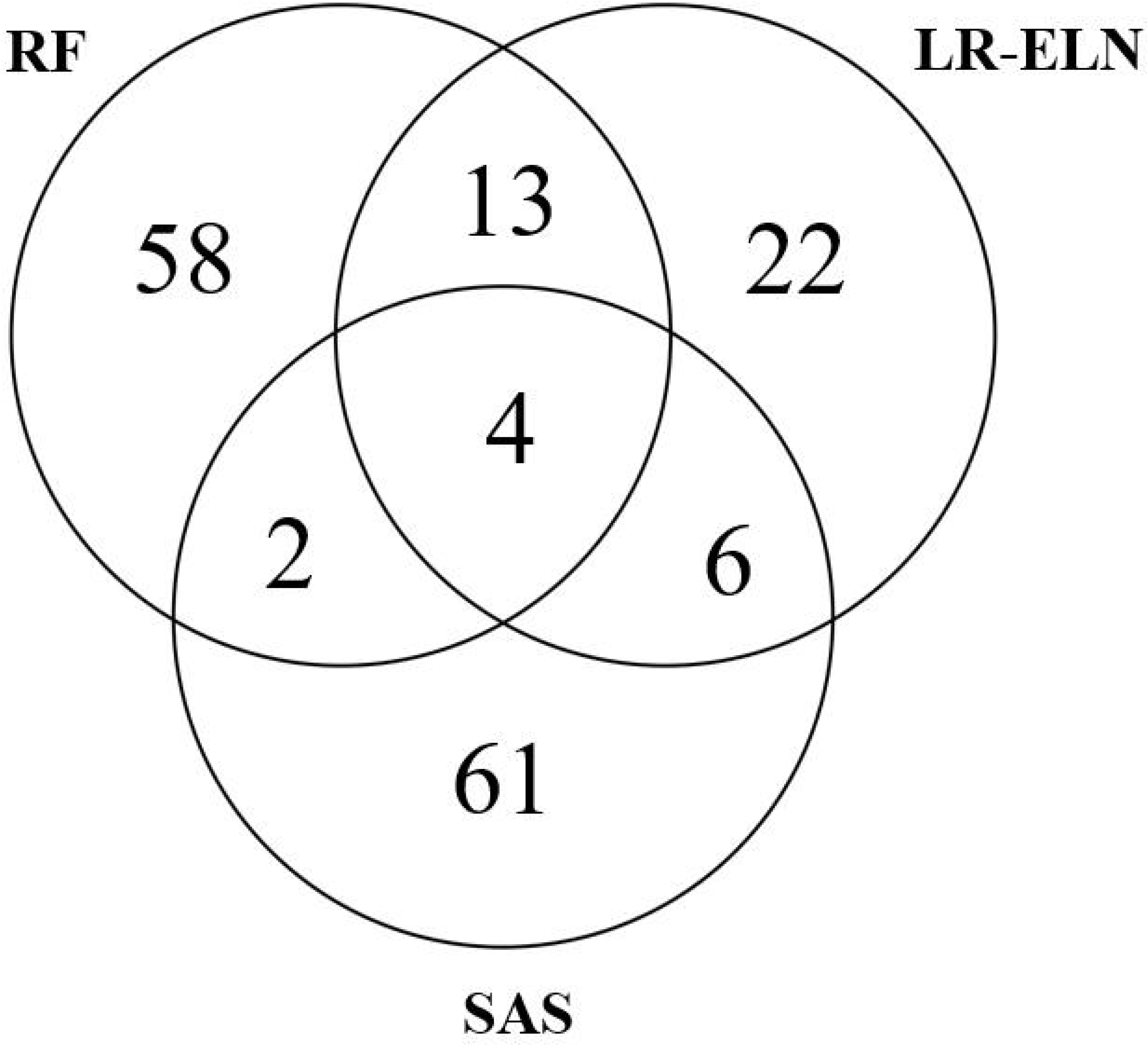
Features selected by the stepwise additive selector, random forest classifier, and elastic net. Venn diagram of features selected by each method. SAS – stepwise additive selector, RF – random forest, LR-ELN – elastic net logistic regression.

**Table 4.**
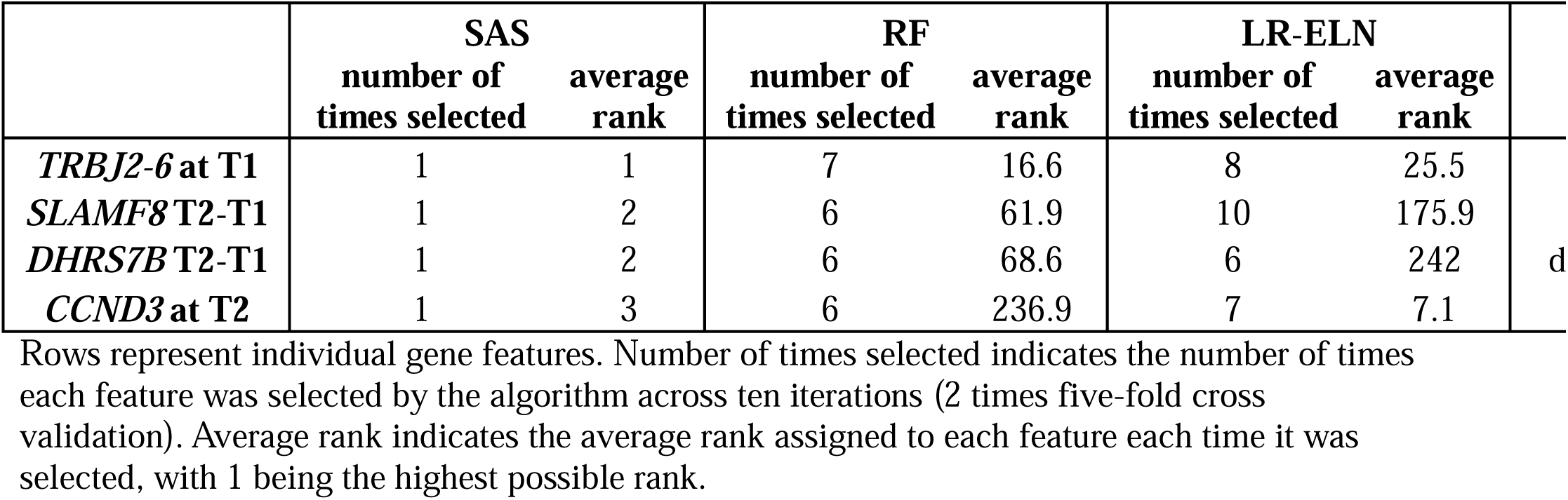
Common features identified by three feature selection approaches.

### Model performance for prediction of sPTB

All models showed promising performance in the training set, with the top performing model was the random forest classifier (AUROC=0.99) (Figure 4, Table 5). However, all five learning models showed significant degradation in performance when applied to the Calgary cohort test set (AUROC range 0.54-0.59) and further degraded when validated externally (AUROC range 0.50-0.52), which indicates a high degree of overfitting.

**Figure 4.**
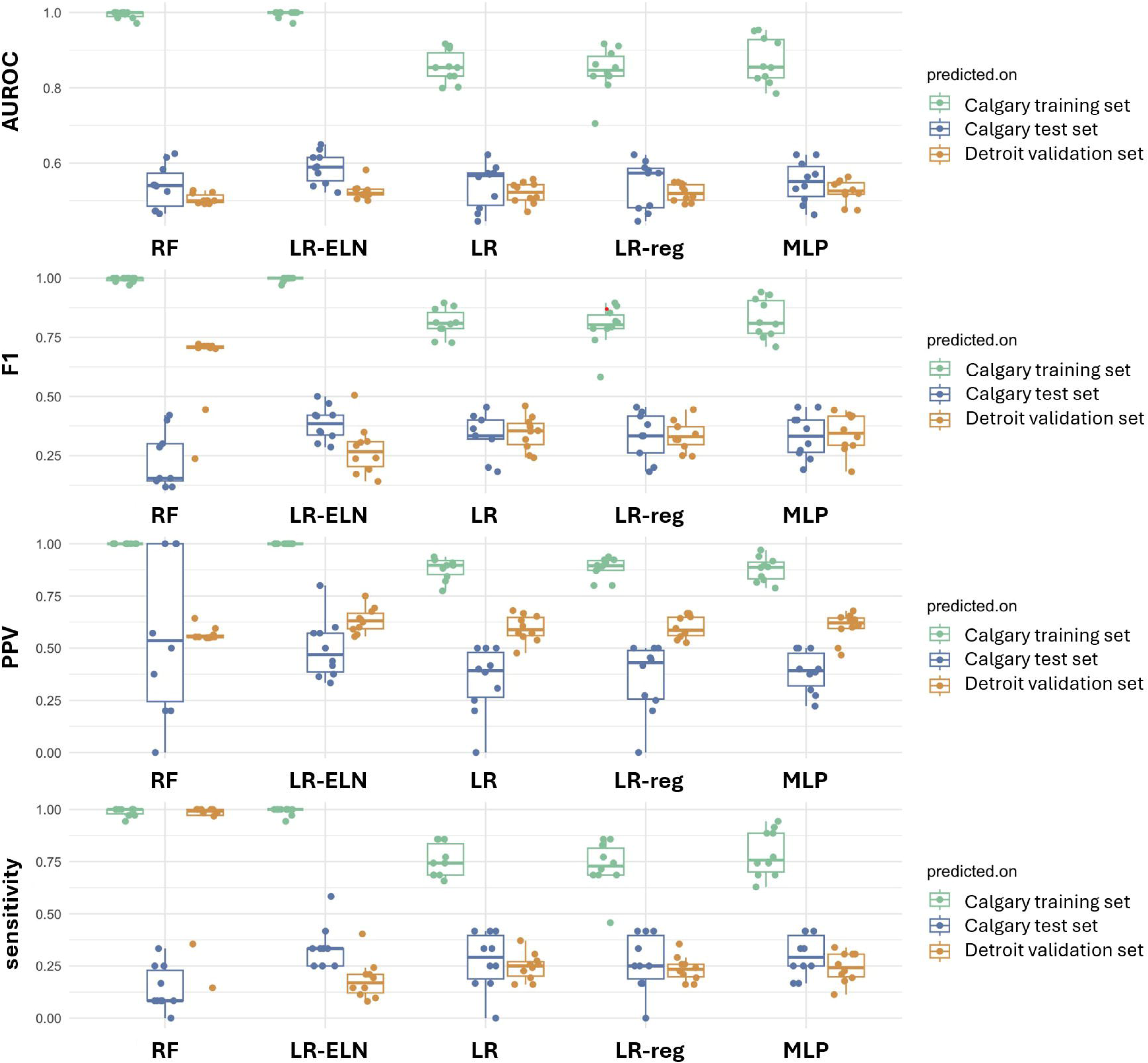
Model performance. Model performance metrics, AUROC (area under the receiver operator curve), F1, PPV (positive predictive value), and sensitivity of each model in the training, internal test and external validation sets.

**Table 5.**
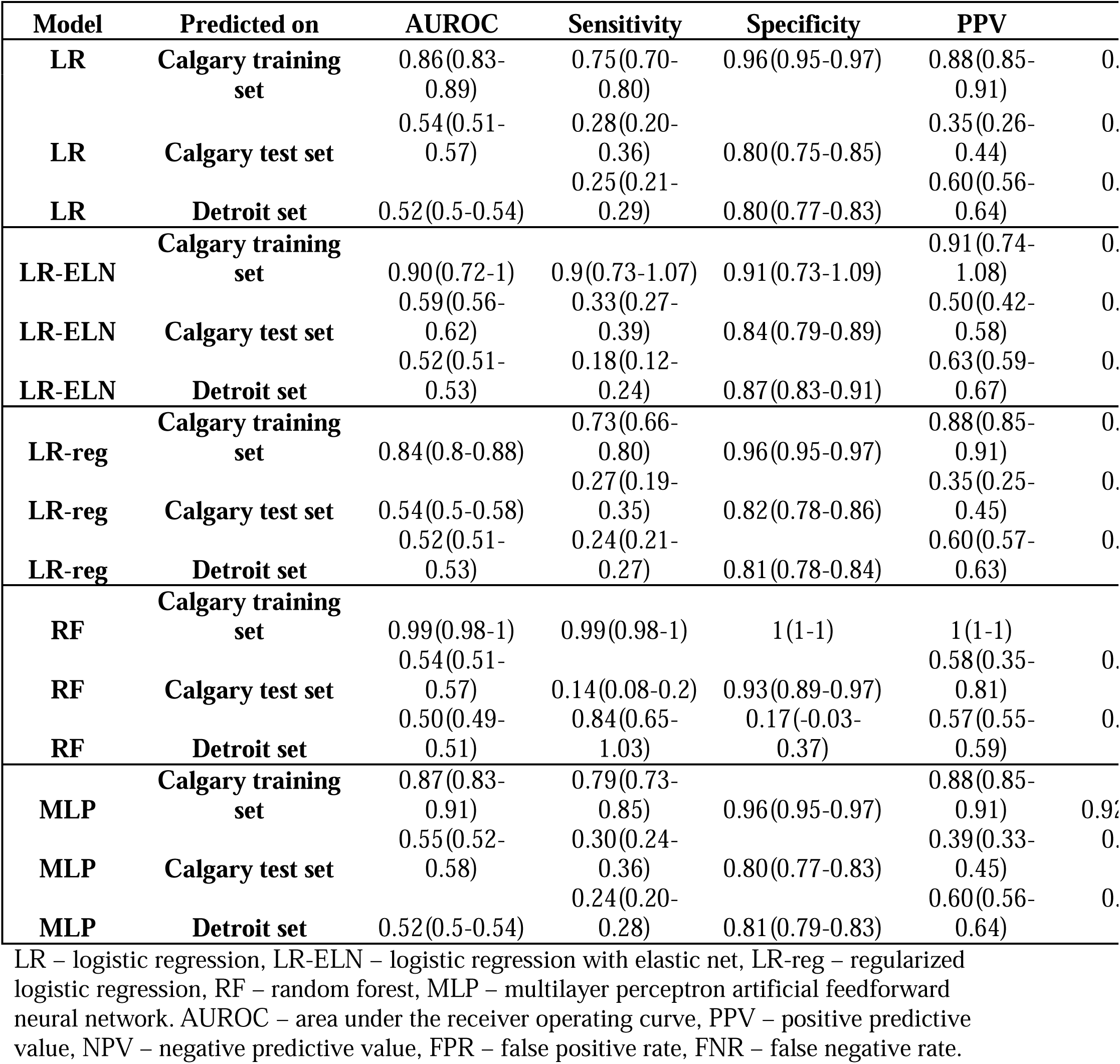
Model performance.

### Assessing overfitting

All models showed significant degradation of performance in both internal and external test sets as compared to training performance, indicating a high degree of overfitting during training. To test the degree of overfitting, the unregularized LR and MLP algorithms were retrained using permuted data, as these represent the lowest and highest complexity models used.

In brief, target labels (sPTB or term) were randomly scrambled during the preprocessing stage to remove any relationship between the ground truth and predictive gene features before proceeding with differential expression analysis, feature selection, and model training as previously described. Using scrambled data, high performance was still observed in the training set, with the highest performance by the MLP algorithm (AUC 0.80). This high performance despite the forced disassociation between true sPTB labels and gene expression suggests overfitting in both these models. Unsurprisingly, model performance was degraded when applied to both the internal and external test sets (Table 6). These results underscore the overfitting we observed for all five machine learning approaches.

**Table 6.** Model performance using permuted data.

| *Model* | *Training Set* | *Test Set* | *Training*<br>*AUROC* | *Test*<br>*AUROC* |
| --- | --- | --- | --- | --- |
| MLP | Calgary | Calgary | 0.80 | 0.49 |
| LR | Calgary | Calgary | 0.75 | 0.50 |
| MLP | Calgary | Detroit | 0.72 | 0.52 |
| LR | Calgary | Detroit | 0.63 | 0.52 |

Model performance as reported by area under the receiver operating curve (AUC) for two prediction algorithms, multilayer perceptron (MLP) and logistic regression (LR) tested internally and validated externally.

## Discussion

We were unable to repeat the findings of Heng et al., [21] to predict spontaneous preterm birth using maternal blood gene expression. Most alarmingly, two of the eight topmost predictive genes were not detectable in blood samples from the same patients, suggesting issues with repeatability in probe-based RNA array methods used here, despite validation of RNA integrity over long-term storage [22]. Indeed, array reliability may be particularly problematic in lowly expressed genes and certain genes may be more subject to poor probe specificity [32, 33], though we were unable to conduct assessment of gene-specific expression levels over long-term storage. Microarray platforms for detection of gene expression signatures may be limited by low specificity of probes, and are not robust to gene variants, leading to inconsistencies in detection which likely impacted reproducibility. Additionally, we were unable to produce a more generalizable model through secondary analysis of the microarray data using alternate learning algorithms, and our results suggest a high degree of overfitting following external validation.

This highlights the importance of repeat and validation studies to meaningfully progress the field of preterm birth prediction.

### Heterogeneity in validation cohorts

Technical and/or biological differences between the training cohort and the cohort used for external validation may have further contributed to the lack of generalizability. One of the strengths of this study was that we were able to identify an external cohort with maternal blood gene expression data collected at the same timepoints during gestation, and used comparable microarray methods for expression analysis, however, batch effects between cohorts may still contribute to the lack of generalizability. Additionally, while the Calgary based training cohort was primarily Caucasian, with a mean age of 31, the population in the external validation cohort from Detroit were mainly African American (92%) and younger (mean age 24). Additionally, the Detroit based cohort had a higher percentage of a prior preterm birth, at 15.5% in the term control population (as compared to 4% in the Calgary cohort) and as high as 50% in the sPTB population (as compared to 17% in the Calgary cohort), suggesting that the Detroit cohort represents a higher risk population with respect to preterm birth. The contribution of the social determinants of health, including systemic racism and unequal access to resources and quality healthcare among different populations including African Americans living in the United States of America, may present distinct mechanisms leading to preterm birth as compared to the population represented in the Calgary cohort.

### Impact of overfitting and data leakage

Reassessment of the prediction model as published by Heng *et al*., [21] suggests that the original model may have been unintentionally impacted by data leakage. Many prediction studies, at least in the field of reproduction, are preceded by observational experiments to explore biomarker patterns as possible predictors and to reduce the number of features for subsequent biomarker discovery. Differential gene expression analysis is frequently used as a filtering step to identify genes associated with an outcome. Often, these observational experiments are conducted on the whole dataset, not the training set. As such, prediction models trained on this data are biased by patterns that exist in the test set. This phenomenon, where information from outside the training dataset is used to create the prediction model, is known as data leakage.

Consequently, the training dataset contains information about the outcome that would not be otherwise available when using the model for prediction, artificially overinflating the predictive performance when the model is applied to the test set. We, the authors, would like to express that we believe data leakage was conducted entirely unintentionally. Indeed, we only identified the data leakage when it was identified by a reviewer in a previous, failed submission for publication, in which we had unintentionally committed the same type of data leakage during our validation study.

Validation with data leakage, in our dataset, demonstrated high predictive performance in the training set (AUC 0.72 with LR, 0.79 with MLP), and was not significantly degraded in the test set (AUC 0.65 with LR, 0.85 with MLP). Note that with the presence of data leakage, the machine learning MLP model had substantially higher performance (AUC 0.85) as compared to analysis without data leakage (AUC 0.54 for comparable dataset). These misleading performance metrics are a result of conducting differential expression analysis prior to splitting the training and test sets. Models were built by preselecting candidate genes identified in the differential expression analysis, which was informed by both the training and test sets, thus, model performance was ultimately biased by information it should not have ‘seen’. This stresses the importance of methodological safeguarding and careful study design, to avoid possible sources of bias and data leakage, particularly in omics or similar datasets that are prone to a high degree of noise. External validation is also a highly powerful tool for testing the generalizability of models that may have been subject to data leakage, and unintentional data leakage likely contributes to the lack of reproducibility in some prediction studies.

One of the primary limitations to prediction using maternal blood gene expression encountered was overfitting and noise within the dataset, which significantly skewed performance estimates. Microarray and similar expression datasets typically contain thousands of features, significantly inflating the feature to observation ratios in this analysis. Feature selection approaches were unable to effectively reduce the noise within this dataset to obtain clinically useful patterns as markers for spontaneous preterm birth. Limited sample size, particularly in the test set, may have impacted our ability to detect true associations. Nonetheless, the sample size of n=165 represents one of the largest maternal blood gene expression datasets available in the pregnant population, a population that is currently understudied. A noisy dataset with high feature to observation ratio likely also contributed to the inability to repeat and/or validate previous findings. Possible consequences of data noise are further exacerbated when using advanced methods such as machine learning, and, as evidenced in the study herein, high complexity/machine learning approaches often do not demonstrate improved predictive performance over traditional methods [34].

### Biological significance of biomarkers identified

We observed limited consistency in genes identified between three feature selection methods, suggesting that their association with sPTB is not robust. Only four common genes were identified by the SAS, elastic net, and random forest classifiers: *TRPJ2-6, SLAMF8, DHRS7B,* and *CCND3.* These identified genes associated with immune responses and cell cycling. The first, *TRPJ2-6,* encodes a protein predicted to be part of the T cell receptor complex in the J region of the variable domain of the T cell receptor beta chain that is involved in antigen recognition [35]. *SLAMF8* encodes a member of the CD2 family of cell surface proteins involved in lymphocyte activation. *DHRS7B* encodes a protein predicted to be involved in lipid biosynthesis, and *CCND3* belongs to the highly conserved cyclin family and regulates the cell cycle during G1/S transition [36]. Taken together, this may suggest that sPTB is associated with changes in immune responses and cell cycling that can be observed in whole blood from the maternal circulation.

### Suggestions for future biomarker discovery

While maternal blood presents an enticing opportunity for minimally invasive prediction, peripheral blood is subject to a high noise signal from various physiological processes occurring within the body unrelated to uterine function during pregnancy. This stresses the need for improved feature identification. Biological compartments including cervicovaginal fluid, amniotic fluid, and the vaginal microbiome may better reflect the physiology of pregnancy and the transition to parturition [37, 38] though sample availability of reproductive and gestational tissues for research purposes is limited. An emerging strategy involves the use of cell-free nucleic acid biomarkers, which can be utilized to identify biomarkers with uterine origin in maternal blood for improved prediction of adverse pregnancy outcomes [39, 40]. Considerable research has been conducted to review the most robust predictors for sPTB, including but not limited to inflammatory biomarkers, maternal characteristics and genetic contributions [41–43], yet the most frequently used risk factors in current literature show variable predictive performance and poor robustness [44]. A recent meta-analysis identified the most robust predictors of PTB, including low gestational weight gain, interpregnancy interval following miscarriage <6months, and sleep-disordered breathing [43], and it is likely that combined biomarker approaches are necessary for prediction [45, 46]. Additionally, current literature often does not distinguish those predictors for a medically indicated PTB from those for sPTB, despite likely distinct aetiologies. It is also worth noting that the pervasive use of convenience sampling in reproductive studies (e.g. secondary analysis of biosamples used for routine antenatal screening) are not necessarily performed proximal to the outcome of interest (PTB). For many subjects, the delay from testing to outcome may make identifying true associations difficult.

### Strategies to improve prediction

Future research in identifying biomarkers for the mechanisms of preterm labour are important and likely requires biologically informed selection over unbiased feature selection, as these methods may suffer more from high noise to feature ratios, especially considering the heterogeneity of PTB and biological variation between individuals. Recent systematic reviews have highlighted that currently described biomarkers have yet to show promising associations for utility in prediction [47]. Yet, recent advancements point to proteomic signatures as a promising biomarker for preterm birth [48], and biomarker discovery targeted to subtypes of PTB such as PPROM [49]. The best models combine an understanding of the features (such as genes, proteins, or patient characteristics) that are most important for determining the outcome and robust methodologies. Identification of robust predictors of sPTB likely requires a combination of advanced selection techniques and an improved understanding of the physiological mechanisms of labour (including known pathways of biomarkers involved in sPTB).

Looking ahead, our recommendations for future research include safeguarding against sources of data leakage, implementing cross-validation techniques as a measure of robustness, external validation as a measurement of overfitting, and prioritizing repeatability and reproducibility of findings. Specific recommendations for addressing data leakage are to ensure testing data has no influence on model training, including in any discovery analysis (such as differential expression analysis) which may precede model training. Dimensionality reduction techniques, such as principal component analysis (PCA), can mediate overfitting by reducing overall dimensionality. However, one disadvantage of dimensionality reduction using techniques like PCA is that they collapse predictors into groups of predictors, which is less enticing for clinical application, as cost and feasibility of clinical testing scales with the number of predictors. Other recommendations for addressing overfitting are to conduct permutation tests to distinguish true patterns from noise in addition to external validation. This likely includes incentivizing repeated studies in published literature and improving data management, storage, and sharing infrastructure, particularly for larger, multicentre studies [10, 11].

### Limitations

The strength of this study was to undertake a replication and external validation, which is, unfortunately, lacking in many prediction studies for preterm birth. However, this study is not without limitations. The current assay was unable to measure miRNA, preventing the measurement of one biomarker *MIR3691*, further, two of the biomarkers were not found to be expressed on repetition of the study. Together this impacted our ability to fully replicate the prediction model . As commercially available microarray assays were used, it leads to important considerations of assay reliability. Additionally, the biomarker *SH3PXD2B* was expressed below the limit of detection in 22% of the samples, for which values were imputed at half the limit of detection. Replacing values with half the limit of the detection has not been shown to significantly impact means when the percentage of imputed values is low, less than 25% [50].

Nonetheless, imputation of missing values can contribute to bias within the measurement, which may explain why were unable to repeat the finding that *SH3PXD2B* expression is different in the preterm population compared to term.

A nested 2:1 case control study design was selected to address class imbalance, as the typical rate of sPTB is ∼10%. However, given the moderate class imbalance in this study design, and the possible bias inherent to resampling or weighting methods, no additional methods to address class imbalance was conducted. There is no consensus on what degree of class imbalance is acceptable, though moderate class imbalances (e.g. 2:1) would have more limited impacts on model training as compared to 10:1. However, it has been suggested error rates for moderate class imbalances (between 1:1 and 1:3) are underestimated, and more problematic than commonly acknowledged [51]. As such, future work in exploring prediction models for preterm birth could consider class weighting or resampling techniques, such as SMOTE (synthetic minority over-sampling) of preterm birth cases or under-sampling of term cases to address this class imbalance, in addition to a case control study design.

## Conclusion

Our study serves as a cautionary tale for researchers, emphasizing the need for transparency, rigorous methodological standards, as well as not only repeating results but validation in external cohorts to advance the field of spontaneous preterm birth prediction responsibly. Our findings also underscore the broader implications for omics studies for discovery analysis, where high feature-to-observation ratios are common, which exacerbates the challenge for mitigating bias and ensuring the reliability of predictive models. For example, differential expression analysis without appropriate training and testing sets for validation introduces inherent bias and limits the generalizability of patterns identified. Testing on internal test sets alone is insufficient for measuring generalizability, especially in the instance of data leakage. Yet, assessments of overfitting and external validation are not standard practice in preterm birth prediction, and the authors stress their importance for meaningful future work in this field.

Current studies on the prediction of sPTB suffer from a lack of external validation, and perhaps unintentional data leakage, leading to a lack of generalized models that would be clinically useful. Recent publication studies have identified various potential predictors of sPTB in both maternal and fetal compartments, though many have not demonstrated robust prediction in external validation nor are repeat studies of promising predictors are not common [52–55].

Further, our results suggest that maternal blood gene expression may either not be predictive of sPTB, or that a high degree of noise limits our ability to detect true associations, stressing the need for a more robust biomarker discovery. Identification of a few potential cell cycling and immune biomarkers presents novel areas for future biomarker discovery. A better understanding of biomarkers associated with sPTB would not only contribute to insight on the mechanism of sPTB, but also to improved prediction of sPTB, allowing for targeted intervention for those at risk for poor maternal and neonatal health outcomes related to prematurity.

## Data Availability

The study involves a secondary analysis of two publicly available gene expression data sets (accession numbers: GSE59491, GSE149440). All other data are made available within the manuscript body and supporting information.

https://www.ncbi.nlm.nih.gov/geo/query/acc.cgi?acc=GSE149440

https://www.ncbi.nlm.nih.gov/geo/query/acc.cgi?acc=GSE59491

## Supporting Information

**S1 Table. Raw fluorescence index and sequencing reads for predictive genes tested in maternal blood.** Isolated RNA from whole maternal blood was analyzed for gene expression using a QuantiGene Plex custom assay (Qiagen) (S1.1) and using bulk RNA sequencing (S1.2).

**S2 Tables. Differential expression analysis results.** Fold differences in each comparison, average expression, F value, p-value and adjusted p-value for each gene identified as differentially expressed in each fold of 10-fold cross validation.

**S3 Table. Feature selection.** Features selected by the stepwise additive selector (SAS), elastic net regression model (LR-ELN) and random forest classifier.

