## Supplementary material for "Machine learning for the prediction of spontaneous preterm birth using early second and third trimester maternal blood gene expression: A Cautionary Tale": S1 Table

**Table S1.1 Raw fluorescence index for predictive genes tested in maternal blood**

|  |  |  | **PROBE-BASED ASSAY** | | | | | | |
| --- | --- | --- | --- | --- | --- | --- | --- | --- | --- |
| **Patient ID** | **Timepoint** | **Group** | **ACAP2** | **LOC101927441** | **ZNF324** | **CST13P** | **LMLN2** | **SH3PXD2B** | **TBX21** |
| **810384** | **T1** | **term** | 10521 | 448 | 561 | <LOD | <LOD | <LOD | 1230 |
| **810384** | **T2** | **term** | 13054 | 259 | 636 | <LOD | <LOD | <LOD | 2102 |
| **810413** | **T1** | **sPTB** | 8369 | 816 | 446 | <LOD | <LOD | <LOD | 744 |
| **810413** | **T2** | **sPTB** | 7803 | 394 | 308 | <LOD | <LOD | <LOD | 725 |
| **810416** | **T2** | **term** | 7207 | 302 | 231 | <LOD | <LOD | <LOD | 237 |
| **810432** | **T1** | **term** | 12151 | 364 | 491 | <LOD | <LOD | <LOD | 674 |
| **810432** | **T2** | **term** | 11386 | 1189 | 678 | <LOD | <LOD | <LOD | 696 |
| **810439** | **T1** | **term** | 8636 | 597 | 411 | <LOD | <LOD | <LOD | 750 |
| **810439** | **T2** | **term** | 9231 | 341 | 420 | <LOD | <LOD | <LOD | 443 |
| **810447** | **T1** | **term** | 18347 | 546 | 822 | <LOD | <LOD | <LOD | 1750 |
| **810447** | **T2** | **term** | 10612 | 949 | 558 | <LOD | <LOD | <LOD | 685 |
| **810477** | **T1** | **sPTB** | 5808 | 268 | 307 | <LOD | <LOD | <LOD | 509 |
| **810494** | **T1** | **term** | 7550 | 378 | 562 | <LOD | <LOD | <LOD | 899 |
| **810494** | **T2** | **term** | 9529 | 286 | 543 | <LOD | <LOD | <LOD | 805 |
| **810507** | **T2** | **term** | 10643 | 1085 | 595 | <LOD | <LOD | <LOD | 1460 |
| **810516** | **T1** | **term** | 10443 | 779 | 655 | <LOD | <LOD | <LOD | 1195 |
| **810516** | **T2** | **term** | 7997 | 354 | 272 | <LOD | <LOD | <LOD | 660 |
| **810518** | **T1** | **term** | 10046 | 318 | 405 | <LOD | <LOD | <LOD | 744 |
| **810519** | **T1** | **term** | 9409 | 759 | 655 | <LOD | <LOD | <LOD | 1324 |
| **810519** | **T2** | **term** | 17148 | 594 | 611 | <LOD | <LOD | 55 | 1109 |
| **810521** | **T1** | **term** | 13232 | 714 | 756 | <LOD | <LOD | <LOD | 836 |
| **810521** | **T2** | **term** | 11215 | 315 | 356 | <LOD | <LOD | <LOD | 518 |
| **810529** | **T1** | **sPTB** | 7394 | 713 | 427 | <LOD | <LOD | <LOD | 1148 |
| **810529** | **T2** | **sPTB** | 8125 | 475 | 394 | <LOD | <LOD | <LOD | 1209 |
| **810567** | **T1** | **term** | 12147 | 519 | 806 | <LOD | <LOD | <LOD | 1040 |
| **810567** | **T2** | **term** | 20533 | 704 | 854 | <LOD | <LOD | 80 | 1420 |
| **810568** | **T1** | **term** | 9065 | 906 | 458 | <LOD | <LOD | <LOD | 1148 |
| **810568** | **T2** | **term** | 10219 | 611 | 514 | <LOD | <LOD | <LOD | 1200 |
| **812231** | **T1** | **term** | 11015 | 387 | 386 | <LOD | <LOD | <LOD | 807 |
| **812231** | **T2** | **term** | 2000 | 39 | 71 | <LOD | <LOD | <LOD | 216 |
| **812234** | **T1** | **term** | 8740 | 328 | 344 | 39 | <LOD | 57 | 661 |
| **812234** | **T2** | **term** | 13045 | 375 | 649 | <LOD | <LOD | 34 | 863 |
| **812268** | **T1** | **term** | 15659 | 962 | 837 | <LOD | 38 | 128 | 2003 |
| **812268** | **T2** | **term** | 10293 | 463 | 561 | <LOD | <LOD | 13 | 1673 |
| **812285** | **T1** | **sPTB** | 9721 | 559 | 356 | <LOD | <LOD | 25 | 516 |
| **812285** | **T2** | **sPTB** | 21953 | 691 | 942 | <LOD | <LOD | 131 | 1130 |
| **812293** | **T1** | **term** | 6838 | 511 | 351 | <LOD | <LOD | <LOD | 536 |
| **812293** | **T2** | **term** | 7889 | 372 | 350 | <LOD | <LOD | 15 | 846 |
| **812302** | **T1** | **sPTB** | 11653 | 551 | 560 | <LOD | <LOD | 35 | 796 |
| **812302** | **T2** | **sPTB** | 14369 | 403 | 614 | <LOD | <LOD | 83 | 685 |
| **812324** | **T1** | **term** | 12207 | 818 | 512 | <LOD | <LOD | 42 | 860 |
| **812324** | **T2** | **term** | 9193 | 360 | 267 | <LOD | <LOD | 43 | 477 |
| **812328** | **T1** | **sPTB** | 5161 | 237 | 209 | <LOD | <LOD | 15 | 404 |
| **812328** | **T2** | **sPTB** | 10315 | 428 | 297 | <LOD | <LOD | 43 | 688 |
| **812342** | **T1** | **sPTB** | 4891 | 244 | 266 | <LOD | <LOD | 14 | 500 |
| **812342** | **T2** | **sPTB** | 5325 | 187 | 312 | <LOD | <LOD | 15 | 752 |
| **812359** | **T1** | **sPTB** | 6951 | 340 | 280 | <LOD | <LOD | 27 | 815 |
| **812359** | **T2** | **sPTB** | 7979 | 316 | 385 | <LOD | <LOD | 43 | 888 |
| **812396** | **T2** | **term** | 14877 | 669 | 646 | 23 | <LOD | 61 | 1650 |
| **812409** | **T1** | **sPTB** | 13873 | 662 | 745 | <LOD | <LOD | 38 | 1664 |
| **812409** | **T2** | **sPTB** | 10574 | 579 | 487 | <LOD | <LOD | 36 | 1196 |
| **812448** | **T1** | **sPTB** | 10080 | 394 | 470 | <LOD | <LOD | 26 | 474 |
| **812459** | **T1** | **sPTB** | 12774 | 713 | 523 | <LOD | <LOD | 43 | 1669 |
| **812459** | **T2** | **sPTB** | 19477 | 933 | 848 | 16 | <LOD | 51 | 2550 |
| **812477** | **T1** | **term** | 11925 | 467 | 446 | <LOD | <LOD | 46 | 927 |
| **812477** | **T2** | **term** | 10516 | 341 | 392 | 19 | <LOD | 27 | 672 |
| **812555** | **T1** | **sPTB** | 8030 | 283 | 268 | <LOD | <LOD | 29 | 631 |
| **812566** | **T1** | **sPTB** | 8179 | 194 | 214 | <LOD | <LOD | 16 | 980 |
| **812566** | **T2** | **sPTB** | 6332 | 175 | 179 | <LOD | <LOD | <LOD | 777 |
| **812573** | **T1** | **term** | 7619 | 234 | 357 | <LOD | <LOD | 25 | 1046 |
| **812573** | **T2** | **term** | 9308 | 446 | 432 | <LOD | <LOD | 23 | 1284 |
| **812574** | **T1** | **term** | 15604 | 765 | 628 | 8 | 4 | 29 | 1231 |
| **812574** | **T2** | **term** | 15903 | 712 | 875 | <LOD | 7 | 85 | 1717 |
| **812587** | **T1** | **sPTB** | 8346 | 403 | 440 | <LOD | 2 | 6 | 1195 |
| **812587** | **T2** | **sPTB** | 18868 | 874 | 802 | 19 | 24 | 19 | 1709 |
| **812590** | **T1** | **term** | 11630 | 511 | 383 | 8 | 7 | 25 | 684 |
| **812590** | **T2** | **term** | 8977 | 381 | 319 | <LOD | 3 | 20 | 487 |
| **815073** | **T1** | **sPTB** | 8923 | 372 | 433 | 6 | 3 | 23 | 649 |
| **815073** | **T2** | **sPTB** | 8547 | 374 | 515 | 8 | 2 | 28 | 638 |
| **815076** | **T1** | **sPTB** | 14440 | 559 | 619 | 6 | 5 | 23 | 714 |
| **815076** | **T2** | **sPTB** | 15865 | 640 | 911 | 9 | 15 | 40 | 1457 |
| **815102** | **T1** | **term** | 9145 | 365 | 466 | <LOD | 1 | 10 | 1301 |
| **815102** | **T2** | **term** | 8960 | 421 | 397 | 7 | 5 | 10 | 720 |
| **815116** | **T1** | **sPTB** | 10645 | 365 | 382 | 8 | 2 | 26 | 584 |
| **815116** | **T2** | **sPTB** | 27778 | 1013 | 818 | 12 | 13 | 208 | 1262 |
| **815137** | **T2** | **term** | 12230 | 442 | 692 | <LOD | 0 | 30 | 1677 |
| **815149** | **T1** | **sPTB** | 11320 | 389 | 440 | 7 | 5 | 31 | 931 |
| **815149** | **T2** | **sPTB** | 8652 | 243 | 270 | 7 | 4 | 37 | 865 |
| **815159** | **T1** | **sPTB** | 10615 | 531 | 764 | <LOD | 1 | 8 | 2078 |
| **815159** | **T2** | **sPTB** | 15715 | 702 | 748 | 9 | 6 | 16 | 2584 |
| **815179** | **T1** | **sPTB** | 16084 | 768 | 758 | 10 | 6 | 73 | 1493 |
| **815179** | **T2** | **sPTB** | 24947 | 917 | 765 | 8 | <LOD | 167 | 1145 |
| **815200** | **T1** | **sPTB** | 7024 | 149 | 346 | <LOD | <LOD | 15 | 905 |
| **815200** | **T2** | **sPTB** | 9396 | 261 | 471 | <LOD | 6 | 10 | 1717 |
| **815218** | **T1** | **sPTB** | 11864 | 369 | 522 | 8 | 1 | 27 | 909 |
| **815218** | **T2** | **sPTB** | 27152 | 659 | 937 | <LOD | 2 | 49 | 1837 |
| **815219** | **T1** | **term** | 11470 | 278 | 498 | <LOD | <LOD | <LOD | 734 |
| **815219** | **T2** | **term** | 10803 | 301 | 351 | 7 | 0 | <LOD | 610 |
| **818023** | **T1** | **sPTB** | 14568 | 590 | 662 | 7 | 2 | 9 | 1466 |
| **818023** | **T2** | **sPTB** | 14248 | 465 | 517 | <LOD | <LOD | 14 | 751 |
| **818025** | **T1** | **term** | 17104 | 771 | 562 | 11 | 11 | 61 | 1042 |
| **818025** | **T2** | **term** | 9931 | 371 | 373 | 6 | <LOD | 32 | 747 |
| **818032** | **T1** | **term** | 6589 | 313 | 203 | <LOD | <LOD | <LOD | 643 |
| **818032** | **T2** | **term** | 11769 | 576 | 550 | 6 | <LOD | <LOD | 981 |
| **818034** | **T1** | **sPTB** | 8306 | 356 | 412 | <LOD | <LOD | 11 | 830 |
| **818034** | **T2** | **sPTB** | 10232 | 332 | 503 | <LOD | <LOD | 8 | 785 |
| **818036** | **T1** | **term** | 19302 | 1143 | 1173 | 17 | 31 | 104 | 1491 |
| **818036** | **T2** | **term** | 14587 | 766 | 684 | <LOD | 10 | 63 | 1193 |
| **818070** | **T1** | **term** | 15070 | 661 | 678 | <LOD | 10 | 32 | 1598 |
| **818070** | **T2** | **term** | 11266 | 389 | 516 | <LOD | <LOD | 24 | 1138 |
| **818080** | **T1** | **term** | 9646 | 431 | 562 | <LOD | 13 | 33 | 1175 |
| **818080** | **T2** | **term** | 8987 | 357 | 593 | <LOD | <LOD | 61 | 853 |
| **818088** | **T1** | **term** | 6352 | 231 | 252 | <LOD | <LOD | 8 | 807 |
| **818088** | **T2** | **term** | 8271 | 259 | 270 | <LOD | <LOD | 33 | 529 |
| **818125** | **T2** | **term** | 6390 | 336 | 265 | <LOD | <LOD | 12 | 664 |
| **818153** | **T1** | **term** | 9096 | 242 | 432 | <LOD | <LOD | 12 | 1091 |
| **818153** | **T2** | **term** | <LOD | <LOD | <LOD | <LOD | <LOD | <LOD | <LOD |
| **818162** | **T1** | **sPTB** | 9659 | 310 | 429 | <LOD | <LOD | 8 | 714 |
| **818162** | **T2** | **sPTB** | 8918 | 576 | 389 | 8 | <LOD | 16 | 678 |
| **818172** | **T1** | **term** | 11342 | 669 | 674 | <LOD | <LOD | 14 | 1797 |
| **818172** | **T2** | **term** | 6049 | 178 | 263 | <LOD | <LOD | <LOD | 824 |
| **818195** | **T2** | **sPTB** | 10956 | 369 | 668 | <LOD | <LOD | 45 | 1054 |
| **818224** | **T1** | **sPTB** | 5882 | 191 | 195 | <LOD | <LOD | 12 | 389 |
| **818224** | **T2** | **sPTB** | 5768 | 186 | 175 | <LOD | <LOD | <LOD | 252 |
| **818241** | **T1** | **sPTB** | 13150 | 582 | 528 | <LOD | <LOD | 18 | 716 |
| **818241** | **T2** | **sPTB** | 13782 | 494 | 556 | <LOD | <LOD | 19 | 703 |
| **818246** | **T1** | **term** | 11693 | 495 | 519 | <LOD | <LOD | 73 | 1030 |
| **818246** | **T2** | **term** | 9909 | 442 | 401 | <LOD | <LOD | 9 | 818 |
| **818249** | **T2** | **sPTB** | 5805 | 201 | 348 | <LOD | <LOD | 19 | 692 |
| **818368** | **T1** | **sPTB** | 11161 | 326 | 405 | 11 | <LOD | 31 | 1130 |
| **818368** | **T2** | **sPTB** | 12116 | 656 | 386 | <LOD | 14 | 23 | 1385 |
| **818409** | **T1** | **sPTB** | 13584 | 577 | 709 | 8 | 12 | 32 | 1360 |
| **818409** | **T2** | **sPTB** | 18292 | 1479 | 1063 | 18 | 31 | 81 | 1549 |
| **818615** | **T1** | **sPTB** | 9118 | 421 | 441 | <LOD | 15 | 29 | 643 |
| **818615** | **T2** | **sPTB** | 9708 | 491 | 570 | <LOD | 16 | 45 | 823 |
| **818626** | **T1** | **sPTB** | 13073 | 566 | 514 | 10 | 18 | 25 | 905 |
| **818670** | **T1** | **sPTB** | 5339 | 309 | 172 | <LOD | 11 | <LOD | 510 |
| **818670** | **T2** | **sPTB** | 5432 | 295 | 220 | <LOD | 13 | 8 | 424 |
| **818684** | **T1** | **sPTB** | 10740 | 533 | 678 | <LOD | 21 | 23 | 990 |
| **818684** | **T2** | **sPTB** | 11040 | 429 | 450 | 8 | 16 | 49 | 800 |
| **818781** | **T1** | **sPTB** | 15625 | 856 | 647 | <LOD | 11 | 31 | 717 |
| **818781** | **T2** | **sPTB** | 8024 | 396 | 349 | 9 | 10 | 19 | 578 |
| **818827** | **T1** | **sPTB** | 6717 | 273 | 253 | <LOD | 13 | 12 | 313 |
| **818827** | **T2** | **sPTB** | 13759 | 580 | 735 | <LOD | 13 | 43 | 896 |
| **830356** | **T1** | **term** | 11857 | 470 | 564 | 11 | 15 | 65 | 808 |
| **830356** | **T2** | **term** | 9362 | 353 | 397 | <LOD | 19 | 21 | 689 |
| **830381** | **T1** | **term** | 12574 | 613 | 602 | <LOD | 23 | 35 | 1293 |
| **830381** | **T2** | **term** | 17513 | 715 | 793 | 11 | 17 | 66 | 1718 |
| **830398** | **T1** | **sPTB** | 7880 | 326 | 376 | <LOD | <LOD | 15 | 530 |
| **830398** | **T2** | **sPTB** | 5742 | 222 | 266 | <LOD | <LOD | 11 | 379 |
| **830401** | **T1** | **term** | 14044 | 731 | 722 | 11 | 20 | 36 | 1855 |
| **830401** | **T2** | **term** | 8420 | 354 | 494 | 10 | 17 | 15 | 1519 |
| **830432** | **T1** | **term** | 11511 | 541 | 597 | <LOD | 14 | 15 | 888 |
| **830432** | **T2** | **term** | 12542 | 553 | 626 | 8 | 14 | 14 | 1737 |
| **830446** | **T1** | **sPTB** | 13968 | 754 | 750 | 13 | 24 | 32 | 2077 |
| **830446** | **T2** | **sPTB** | 9789 | 363 | 312 | <LOD | 8 | 31 | 778 |
| **830505** | **T1** | **sPTB** | 5069 | 179 | 182 | <LOD | <LOD | 20 | 432 |
| **830518** | **T1** | **term** | 9018 | 334 | 486 | 10 | 12 | 15 | 736 |
| **830518** | **T2** | **term** | 7527 | 289 | 313 | <LOD | 12 | 13 | 393 |
| **830533** | **T1** | **term** | 15133 | 545 | 636 | 9 | 19 | 42 | 1405 |
| **830533** | **T2** | **term** | 14853 | 601 | 583 | 16 | 7 | 43 | 1283 |
| **830560** | **T2** | **sPTB** | 10560 | 413 | 728 | <LOD | 8 | 38 | 2622 |
| **830561** | **T1** | **term** | 18163 | 844 | 921 | 11 | 20 | 65 | 2094 |
| **830561** | **T2** | **term** | 10214 | 479 | 504 | 11 | 15 | 56 | 956 |
| **830651** | **T1** | **sPTB** | 6714 | 244 | 388 | 12 | 11 | 18 | 278 |
| **830651** | **T2** | **sPTB** | 6425 | 210 | 281 | 9 | 10 | 15 | 205 |
| **830656** | **T1** | **term** | 11660 | 430 | 676 | <LOD | 5 | 16 | 1190 |
| **830656** | **T2** | **term** | 9742 | 314 | 489 | 10 | 12 | 24 | 1144 |
| **830741** | **T1** | **term** | 12406 | 412 | 815 | <LOD | 11 | 38 | 1810 |
| **830741** | **T2** | **term** | 10484 | 341 | 408 | 9 | 10 | 22 | 1109 |
| **830762** | **T1** | **sPTB** | 12408 | 465 | 530 | 10 | 10 | 22 | 727 |
| **830762** | **T2** | **sPTB** | 6857 | 216 | 338 | 9 | 6 | 16 | 610 |
| **830872** | **T1** | **sPTB** | 9483 | 285 | 499 | 17 | 5 | 22 | 1125 |
| **830872** | **T2** | **sPTB** | 16071 | 583 | 733 | 10 | 11 | 84 | 1399 |
| **830909** | **T1** | **sPTB** | 11761 | 389 | 516 | 9 | 5 | 21 | 2018 |
| **830909** | **T2** | **sPTB** | 12797 | 398 | 602 | 14 | 6 | 39 | 1650 |

**Table S1.2 Normalized reads per gene for predictive genes tested in maternal blood.**

|  |  |  | **RNA SEQUENCING** | | | | | | | |
| --- | --- | --- | --- | --- | --- | --- | --- | --- | --- | --- |
| **Patient ID** | **Timepoint** | **Group** | **ACAP2** | **LOC101927441** | **ZNF324** | **CST13P** | **LMLN2** | **SH3PXD2B** | **TBX21** | **MIR3691** |
| **810384** | **T1** | **term** | 3429 | 50 | 214 | 0 | 2 | 14 | 845 | 0 |
| **810384** | **T2** | **term** | 3909 | 47 | 353 | 0 | 4 | 21 | 2011 | 0 |
| **810413** | **T1** | **sPTB** | 1435 | 16 | 60 | 0 | 0 | 3 | 221 | 0 |
| **810413** | **T2** | **sPTB** | 2116 | 19 | 115 | 0 | 2 | 16 | 393 | 0 |
| **810416** | **T2** | **term** | 2178 | 20 | 126 | 0 | 1 | 31 | 228 | 0 |
| **810432** | **T1** | **term** | 2286 | 46 | 116 | 0 | 0 | 8 | 266 | 0 |
| **810432** | **T2** | **term** | 1934 | 37 | 114 | 0 | 0 | 5 | 300 | 0 |
| **810439** | **T1** | **term** | 4558 | 50 | 287 | 0 | 1 | 12 | 997 | 0 |
| **810439** | **T2** | **term** | 1396 | 13 | 122 | 0 | 2 | 11 | 261 | 0 |
| **810447** | **T1** | **term** | 3470 | 28 | 165 | 0 | 0 | 30 | 489 | 0 |
| **810447** | **T2** | **term** | 5884 | 69 | 491 | 0 | 8 | 64 | 960 | 0 |
| **810477** | **T1** | **sPTB** | 1119 | 12 | 140 | 0 | 1 | 13 | 429 | 0 |
| **810494** | **T1** | **term** | 1996 | 20 | 163 | 0 | 1 | 28 | 585 | 0 |
| **810494** | **T2** | **term** | 2287 | 25 | 156 | 0 | 0 | 23 | 467 | 0 |
| **810507** | **T2** | **term** | 3021 | 31 | 180 | 0 | 1 | 37 | 685 | 0 |
| **810516** | **T1** | **term** | 2269 | 31 | 166 | 0 | 0 | 13 | 567 | 0 |
| **810516** | **T2** | **term** | 1263 | 9 | 73 | 0 | 0 | 12 | 272 | 0 |
| **810518** | **T1** | **term** | 3057 | 39 | 249 | 0 | 1 | 55 | 693 | 0 |
| **810519** | **T1** | **term** | 2338 | 12 | 159 | 0 | 2 | 29 | 592 | 0 |
| **810519** | **T2** | **term** | 1795 | 8 | 96 | 0 | 0 | 28 | 294 | 0 |
| **810521** | **T1** | **term** | 3057 | 32 | 168 | 0 | 1 | 25 | 494 | 0 |
| **810521** | **T2** | **term** | 1595 | 15 | 110 | 0 | 2 | 30 | 225 | 0 |
| **810529** | **T1** | **sPTB** | 1698 | 38 | 145 | 0 | 0 | 28 | 813 | 0 |
| **810529** | **T2** | **sPTB** | 1837 | 36 | 152 | 0 | 1 | 16 | 824 | 0 |
| **810567** | **T1** | **term** | 3002 | 28 | 256 | 0 | 4 | 29 | 569 | 0 |
| **810567** | **T2** | **term** | 4693 | 30 | 263 | 0 | 3 | 73 | 606 | 0 |
| **810568** | **T1** | **term** | 2812 | 47 | 217 | 0 | 0 | 27 | 892 | 0 |
| **810568** | **T2** | **term** | 3556 | 57 | 293 | 0 | 0 | 40 | 1323 | 0 |
| **812231** | **T1** | **term** | 2575 | 22 | 127 | 0 | 0 | 40 | 333 | 1 |
| **812231** | **T2** | **term** | 2898 | 26 | 202 | 0 | 1 | 32 | 611 | 0 |
| **812234** | **T1** | **term** | 2664 | 18 | 138 | 0 | 0 | 14 | 388 | 0 |
| **812234** | **T2** | **term** | 2945 | 33 | 181 | 0 | 1 | 27 | 504 | 0 |
| **812268** | **T1** | **term** | 1655 | 24 | 153 | 0 | 0 | 25 | 703 | 0 |
| **812268** | **T2** | **term** | 3685 | 35 | 209 | 0 | 1 | 14 | 999 | 0 |
| **812285** | **T1** | **sPTB** | 4326 | 34 | 199 | 0 | 1 | 57 | 462 | 0 |
| **812285** | **T2** | **sPTB** | 4721 | 56 | 209 | 0 | 2 | 81 | 481 | 0 |
| **812293** | **T1** | **term** | 1445 | 6 | 102 | 0 | 3 | 13 | 228 | 0 |
| **812293** | **T2** | **term** | 1139 | 6 | 91 | 0 | 1 | 12 | 277 | 0 |
| **812302** | **T1** | **sPTB** | 1908 | 15 | 141 | 0 | 1 | 23 | 363 | 0 |
| **812302** | **T2** | **sPTB** | 1875 | 15 | 164 | 0 | 1 | 39 | 255 | 0 |
| **812324** | **T1** | **term** | 3134 | 41 | 148 | 0 | 1 | 35 | 391 | 0 |
| **812324** | **T2** | **term** | 2195 | 12 | 117 | 0 | 0 | 33 | 240 | 0 |
| **812328** | **T1** | **sPTB** | 793 | 5 | 47 | 0 | 1 | 10 | 143 | 0 |
| **812328** | **T2** | **sPTB** | 2432 | 32 | 91 | 0 | 0 | 42 | 390 | 0 |
| **812342** | **T1** | **sPTB** | 1260 | 15 | 121 | 0 | 2 | 13 | 342 | 0 |
| **812342** | **T2** | **sPTB** | 1159 | 18 | 120 | 0 | 0 | 7 | 413 | 0 |
| **812359** | **T1** | **sPTB** | 2273 | 35 | 200 | 0 | 2 | 36 | 898 | 0 |
| **812359** | **T2** | **sPTB** | 2114 | 31 | 190 | 0 | 2 | 28 | 842 | 0 |
| **812396** | **T2** | **term** | 2825 | 32 | 241 | 0 | 4 | 55 | 975 | 0 |
| **812409** | **T1** | **sPTB** | 3116 | 58 | 230 | 0 | 3 | 36 | 974 | 0 |
| **812409** | **T2** | **sPTB** | 2319 | 46 | 183 | 0 | 0 | 27 | 790 | 0 |
| **812448** | **T1** | **sPTB** | 3465 | 35 | 206 | 0 | 1 | 14 | 351 | 0 |
| **812459** | **T1** | **sPTB** | 3783 | 28 | 217 | 0 | 0 | 44 | 981 | 0 |
| **812459** | **T2** | **sPTB** | 5218 | 52 | 298 | 0 | 0 | 42 | 1472 | 0 |
| **812477** | **T1** | **term** | 2303 | 28 | 192 | 0 | 0 | 48 | 642 | 1 |
| **812477** | **T2** | **term** | 1867 | 29 | 137 | 0 | 0 | 19 | 452 | 0 |
| **812555** | **T1** | **sPTB** | 2047 | 20 | 104 | 0 | 1 | 14 | 329 | 0 |
| **812566** | **T1** | **sPTB** | 2076 | 10 | 106 | 0 | 0 | 12 | 485 | 0 |
| **812566** | **T2** | **sPTB** | 2493 | 20 | 138 | 0 | 1 | 6 | 569 | 0 |
| **812573** | **T1** | **term** | 1883 | 15 | 145 | 0 | 1 | 6 | 538 | 0 |
| **812573** | **T2** | **term** | 2033 | 9 | 164 | 0 | 3 | 19 | 596 | 0 |
| **812574** | **T1** | **term** | 2622 | 23 | 148 | 0 | 0 | 24 | 418 | 0 |
| **812574** | **T2** | **term** | 4297 | 44 | 251 | 0 | 0 | 71 | 819 | 0 |
| **812587** | **T1** | **sPTB** | 2395 | 14 | 136 | 0 | 1 | 14 | 582 | 0 |
| **812587** | **T2** | **sPTB** | 1653 | 23 | 160 | 0 | 1 | 11 | 415 | 0 |
| **812590** | **T1** | **term** | 2391 | 26 | 110 | 0 | 4 | 29 | 265 | 0 |
| **812590** | **T2** | **term** | 2153 | 29 | 112 | 0 | 1 | 26 | 258 | 0 |
| **815073** | **T1** | **sPTB** | 1923 | 19 | 139 | 0 | 1 | 34 | 369 | 0 |
| **815073** | **T2** | **sPTB** | 2205 | 28 | 182 | 0 | 0 | 42 | 543 | 0 |
| **815076** | **T1** | **sPTB** | 3057 | 20 | 163 | 0 | 0 | 29 | 419 | 0 |
| **815076** | **T2** | **sPTB** | 4734 | 55 | 264 | 0 | 7 | 51 | 770 | 0 |
| **815102** | **T1** | **term** | 2332 | 30 | 182 | 0 | 0 | 13 | 690 | 0 |
| **815102** | **T2** | **term** | 3148 | 34 | 176 | 0 | 2 | 31 | 462 | 0 |
| **815116** | **T1** | **sPTB** | 2523 | 35 | 107 | 0 | 1 | 21 | 294 | 0 |
| **815116** | **T2** | **sPTB** | 3524 | 39 | 164 | 0 | 1 | 95 | 311 | 0 |
| **815137** | **T2** | **term** | 2877 | 18 | 268 | 0 | 0 | 56 | 948 | 0 |
| **815149** | **T1** | **sPTB** | 1469 | 14 | 94 | 0 | 1 | 26 | 313 | 0 |
| **815149** | **T2** | **sPTB** | 1379 | 19 | 127 | 0 | 0 | 33 | 482 | 0 |
| **815159** | **T1** | **sPTB** | 4618 | 43 | 333 | 0 | 0 | 23 | 1534 | 0 |
| **815159** | **T2** | **sPTB** | 4105 | 45 | 250 | 0 | 0 | 27 | 1395 | 0 |
| **815179** | **T1** | **sPTB** | 4619 | 51 | 277 | 0 | 0 | 84 | 881 | 0 |
| **815179** | **T2** | **sPTB** | 6123 | 69 | 288 | 0 | 1 | 125 | 649 | 0 |
| **815200** | **T1** | **sPTB** | 1122 | 14 | 103 | 0 | 3 | 10 | 483 | 0 |
| **815200** | **T2** | **sPTB** | 1132 | 15 | 126 | 0 | 2 | 13 | 659 | 0 |
| **815218** | **T1** | **sPTB** | 1952 | 12 | 156 | 0 | 3 | 17 | 393 | 0 |
| **815218** | **T2** | **sPTB** | 3091 | 27 | 199 | 0 | 0 | 40 | 804 | 0 |
| **815219** | **T1** | **term** | 3060 | 27 | 198 | 0 | 2 | 9 | 538 | 0 |
| **815219** | **T2** | **term** | 1963 | 15 | 166 | 0 | 1 | 5 | 485 | 0 |
| **818023** | **T1** | **sPTB** | 2728 | 25 | 156 | 0 | 0 | 11 | 579 | 0 |
| **818023** | **T2** | **sPTB** | 3498 | 20 | 145 | 0 | 0 | 3 | 376 | 0 |
| **818025** | **T1** | **term** | 4385 | 23 | 153 | 0 | 0 | 46 | 412 | 0 |
| **818025** | **T2** | **term** | 2310 | 11 | 99 | 0 | 0 | 36 | 329 | 0 |
| **818032** | **T1** | **term** | 2468 | 23 | 137 | 0 | 1 | 10 | 512 | 0 |
| **818032** | **T2** | **term** | 2132 | 22 | 143 | 0 | 2 | 3 | 422 | 0 |
| **818034** | **T1** | **sPTB** | 2941 | 39 | 129 | 0 | 0 | 19 | 530 | 0 |
| **818034** | **T2** | **sPTB** | 1922 | 25 | 110 | 0 | 0 | 10 | 348 | 0 |
| **818036** | **T1** | **term** | 8322 | 124 | 432 | 0 | 1 | 76 | 959 | 0 |
| **818036** | **T2** | **term** | 4378 | 46 | 257 | 0 | 0 | 58 | 576 | 0 |
| **818070** | **T1** | **term** | 2971 | 26 | 217 | 0 | 1 | 22 | 626 | 0 |
| **818070** | **T2** | **term** | 3264 | 37 | 224 | 0 | 1 | 24 | 721 | 2 |
| **818080** | **T1** | **term** | 2371 | 15 | 141 | 0 | 3 | 36 | 548 | 0 |
| **818080** | **T2** | **term** | 1930 | 20 | 154 | 0 | 0 | 35 | 351 | 0 |
| **818088** | **T1** | **term** | 1451 | 23 | 84 | 0 | 0 | 21 | 372 | 0 |
| **818088** | **T2** | **term** | 1955 | 19 | 106 | 0 | 1 | 56 | 262 | 0 |
| **818125** | **T2** | **term** | 2192 | 25 | 128 | 0 | 1 | 22 | 464 | 0 |
| **818153** | **T1** | **term** | 1753 | 24 | 164 | 0 | 0 | 25 | 782 | 0 |
| **818153** | **T2** | **term** | 1353 | 17 | 194 | 0 | 4 | 33 | 1133 | 0 |
| **818162** | **T1** | **sPTB** | 2669 | 10 | 151 | 0 | 1 | 18 | 463 | 0 |
| **818162** | **T2** | **sPTB** | 2396 | 20 | 132 | 0 | 0 | 14 | 405 | 0 |
| **818172** | **T1** | **term** | 3405 | 34 | 254 | 0 | 4 | 24 | 858 | 0 |
| **818172** | **T2** | **term** |  |  |  |  |  |  |  |  |
| **818195** | **T2** | **sPTB** | 1708 | 13 | 152 | 0 | 1 | 43 | 376 | 0 |
| **818224** | **T1** | **sPTB** | 2010 | 20 | 100 | 0 | 0 | 28 | 348 | 0 |
| **818224** | **T2** | **sPTB** | 1323 | 20 | 60 | 0 | 0 | 15 | 170 | 0 |
| **818241** | **T1** | **sPTB** | 3227 | 40 | 134 | 0 | 1 | 21 | 358 | 0 |
| **818241** | **T2** | **sPTB** | 2411 | 34 | 133 | 0 | 0 | 28 | 276 | 0 |
| **818246** | **T1** | **term** | 1457 | 16 | 93 | 0 | 1 | 29 | 293 | 0 |
| **818246** | **T2** | **term** | 1987 | 28 | 108 | 0 | 0 | 14 | 363 | 0 |
| **818249** | **T2** | **sPTB** | 1488 | 10 | 125 | 0 | 2 | 20 | 372 | 0 |
| **818368** | **T1** | **sPTB** | 3517 | 27 | 183 | 0 | 0 | 56 | 862 | 0 |
| **818368** | **T2** | **sPTB** | 2298 | 24 | 140 | 0 | 0 | 24 | 614 | 0 |
| **818409** | **T1** | **sPTB** | 4207 | 43 | 214 | 0 | 3 | 34 | 656 | 0 |
| **818409** | **T2** | **sPTB** | 6284 | 79 | 359 | 0 | 5 | 60 | 1014 | 0 |
| **818615** | **T1** | **sPTB** | 2628 | 19 | 168 | 0 | 0 | 41 | 398 | 0 |
| **818615** | **T2** | **sPTB** | 2169 | 19 | 170 | 0 | 1 | 25 | 467 | 0 |
| **818626** | **T1** | **sPTB** | 2633 | 12 | 150 | 0 | 0 | 26 | 433 | 0 |
| **818670** | **T1** | **sPTB** | 1190 | 25 | 81 | 0 | 0 | 13 | 260 | 0 |
| **818670** | **T2** | **sPTB** | 1389 | 9 | 76 | 0 | 1 | 6 | 236 | 0 |
| **818684** | **T1** | **sPTB** | 4056 | 39 | 294 | 0 | 0 | 20 | 779 | 0 |
| **818684** | **T2** | **sPTB** | 1769 | 21 | 108 | 0 | 0 | 35 | 420 | 0 |
| **818781** | **T1** | **sPTB** | 4600 | 70 | 318 | 0 | 2 | 65 | 549 | 0 |
| **818781** | **T2** | **sPTB** | 2876 | 26 | 168 | 0 | 0 | 41 | 465 | 0 |
| **818827** | **T1** | **sPTB** | 1611 | 30 | 124 | 0 | 0 | 14 | 287 | 0 |
| **818827** | **T2** | **sPTB** | 3444 | 36 | 234 | 0 | 2 | 35 | 578 | 0 |
| **830356** | **T1** | **term** | 1648 | 4 | 141 | 0 | 0 | 34 | 273 | 0 |
| **830356** | **T2** | **term** | 1830 | 14 | 116 | 0 | 2 | 15 | 304 | 0 |
| **830381** | **T1** | **term** | 1959 | 18 | 157 | 0 | 0 | 42 | 426 | 0 |
| **830381** | **T2** | **term** | 3130 | 29 | 163 | 0 | 0 | 30 | 562 | 0 |
| **830398** | **T1** | **sPTB** | 2927 | 32 | 176 | 0 | 1 | 23 | 484 | 0 |
| **830398** | **T2** | **sPTB** | 2746 | 39 | 168 | 0 | 0 | 16 | 522 | 0 |
| **830401** | **T1** | **term** | 3045 | 35 | 180 | 0 | 1 | 32 | 804 | 0 |
| **830401** | **T2** | **term** | 3411 | 47 | 173 | 0 | 3 | 23 | 1188 | 1 |
| **830432** | **T1** | **term** | 3816 | 40 | 218 | 0 | 3 | 10 | 565 | 0 |
| **830432** | **T2** | **term** | 3006 | 25 | 213 | 0 | 1 | 19 | 877 | 0 |
| **830446** | **T1** | **sPTB** | 5279 | 47 | 367 | 0 | 2 | 45 | 1634 | 0 |
| **830446** | **T2** | **sPTB** | 2184 | 23 | 117 | 0 | 1 | 17 | 438 | 0 |
| **830505** | **T1** | **sPTB** | 1298 | 9 | 54 | 0 | 0 | 19 | 218 | 0 |
| **830518** | **T1** | **term** | 2939 | 36 | 237 | 0 | 0 | 15 | 486 | 0 |
| **830518** | **T2** | **term** | 1441 | 11 | 73 | 0 | 0 | 6 | 147 | 0 |
| **830533** | **T1** | **term** | 3233 | 41 | 271 | 0 | 0 | 48 | 962 | 0 |
| **830533** | **T2** | **term** | 3121 | 36 | 317 | 0 | 0 | 69 | 849 | 0 |
| **830560** | **T2** | **sPTB** | 2035 | 50 | 275 | 0 | 0 | 29 | 1609 | 0 |
| **830561** | **T1** | **term** | 4090 | 55 | 258 | 0 | 1 | 37 | 947 | 0 |
| **830561** | **T2** | **term** | 2003 | 21 | 120 | 0 | 2 | 30 | 338 | 0 |
| **830651** | **T1** | **sPTB** | 1733 | 29 | 152 | 0 | 1 | 24 | 184 | 0 |
| **830651** | **T2** | **sPTB** | 1388 | 15 | 112 | 0 | 0 | 10 | 174 | 0 |
| **830656** | **T1** | **term** | 2987 | 24 | 196 | 0 | 1 | 6 | 534 | 0 |
| **830656** | **T2** | **term** | 1937 | 28 | 131 | 0 | 1 | 12 | 480 | 0 |
| **830741** | **T1** | **term** | 3767 | 52 | 352 | 0 | 2 | 26 | 1439 | 0 |
| **830741** | **T2** | **term** | 3093 | 32 | 180 | 0 | 0 | 38 | 688 | 0 |
| **830762** | **T1** | **sPTB** | 2426 | 29 | 129 | 0 | 0 | 14 | 317 | 0 |
| **830762** | **T2** | **sPTB** | 1989 | 32 | 136 | 0 | 0 | 17 | 392 | 0 |
| **830872** | **T1** | **sPTB** | 2495 | 19 | 166 | 0 | 0 | 28 | 675 | 0 |
| **830872** | **T2** | **sPTB** | 2525 | 15 | 146 | 0 | 0 | 40 | 474 | 0 |
| **830909** | **T1** | **sPTB** | 2081 | 24 | 198 | 0 | 1 | 16 | 1265 | 0 |
| **830909** | **T2** | **sPTB** | 2110 | 24 | 156 | 0 | 2 | 32 | 876 | 0 |
