## Supplementary material for "Machine learning for the prediction of spontaneous preterm birth using early second and third trimester maternal blood gene expression: A Cautionary Tale": S3 Table

**S3 Table Feature selection.**

|  |  | **Random forest** | | | **SAS** | | **Elastic net regression** | |  |
| --- | --- | --- | --- | --- | --- | --- | --- | --- | --- |
| **feature** | **# times DE** | **# times selected** | **mean rank** | **mean VIP** | **# times selected** | **mean rank** | **# times selected** | **mean rank** | **gene** |
| **28623_at_T1** | 10 | 7 | 16.6 | 0.501871 | 1 | 1 | 8 | 25.5 | *TRBJ2-6* |
| **25979_at_T1** | 8 | 4 | 26.125 | 0.651527 | 1 | 1 | 7 | 39 | *DHRS7B* |
| **51258_at_T2** | 5 | 5 | 1.4 | 0.124737 | 2 | 1 | 5 | 140 | *MRPL51* |
| **84081_at_T2** | 1 | 1 | 3 | 0.051026 | 1 | 1 | 1 | 50 | *NSRP1* |
| **25966_at_T1** | 1 | 1 | 2 | 0.174166 | 1 | 1 | 1 | 55 | *C2CD2* |
| **83988_at_T1** | 1 | 1 | 46 | 0.152008 | 1 | 1 | 1 | 65 | *NCALD* |
| **57677_at_T2** | 1 | 1 | 7 | 0.313016 | 1 | 1 | 1 | 274 | *ZFP14* |
| **9221_at_T1** | 3 | 2 | 12 | 0.593319 | 1 | 1 | 0 | 0 | *NOLC1* |
| **7695_at_T2** | 2 | 2 | 8 | 0.276011 | 2 | 1.5 | 2 | 96 | *ZNF136* |
| **56833_at_T2-T1** | 10 | 6 | 61.9 | 0.736985 | 1 | 2 | 10 | 175.9 | *SLAMF8* |
| **25979_at_T2-T1** | 8 | 6 | 68.625 | 0.65671 | 1 | 2 | 6 | 242 | *DHRS7B* |
| **122618_at_T1** | 10 | 8 | 135.6 | 0.732013 | 1 | 2 | 4 | 22 | *PLD4* |
| **2887_at_T1** | 9 | 4 | 31.55556 | 0.75549 | 1 | 2 | 4 | 27.25 | *GRB10* |
| **23492_at_T1** | 6 | 3 | 120.1667 | 0.815062 | 1 | 2 | 4 | 46 | *CBX7* |
| **23075_at_T2** | 3 | 2 | 31.66667 | 0.623434 | 1 | 2 | 3 | 96.33333 | *SWAP70* |
| **10980_at_T1** | 2 | 2 | 10.5 | 0.306333 | 1 | 2 | 2 | 55.5 | *COPS6* |
| **11180_at_T2-T1** | 1 | 1 | 2 | 0.179318 | 1 | 2 | 1 | 178 | *WDR6* |
| **826_at_T2** | 4 | 3 | 199.75 | 0.892936 | 1 | 2 | 0 | 0 | *CAPNS1* |
| **896_at_T2** | 10 | 6 | 236.9 | 0.93209 | 1 | 3 | 7 | 147.1429 | *CCND3* |
| **64746_at_T1** | 10 | 6 | 35 | 0.681344 | 1 | 3 | 4 | 21 | *ACBD3* |
| **64805_at_T1** | 10 | 5 | 54.5 | 0.729655 | 1 | 3 | 4 | 18.25 | *P2RY12* |
| **28395_at_T1** | 8 | 5 | 189 | 0.8682 | 1 | 3 | 3 | 75 | *IGHV4-34* |
| **26996_at_T2-T1** | 4 | 2 | 63.25 | 0.871989 | 1 | 3 | 3 | 229.3333 | *GPR160* |
| **9836_at_T1** | 1 | 1 | 10 | 0.281066 | 1 | 3 | 1 | 31 | *LCMT2* |
| **9529_at_T2-T1** | 1 | 1 | 35 | 0.530122 | 1 | 3 | 1 | 201 | *BAG5* |
| **2919_at_T2-T1** | 1 | 1 | 6 | 0.298979 | 1 | 3 | 1 | 223 | *CXCL1* |
| **9159_at_T2-T1** | 3 | 2 | 36.66667 | 0.758389 | 1 | 3 | 0 | 0 | *PCSK7* |
| **80314_at_T2** | 8 | 3 | 24.125 | 0.806199 | 1 | 4 | 6 | 113 | *EPC1* |
| **8655_at_T2** | 8 | 3 | 194.875 | 0.892309 | 1 | 4 | 4 | 156.5 | *DYNLL1* |
| **51582_at_T1** | 10 | 4 | 140.6 | 0.920421 | 1 | 4 | 2 | 18.5 | *AZIN1* |
| **60386_at_T2-T1** | 2 | 2 | 7 | 0.19208 | 1 | 4 | 2 | 169 | *SLC25A19* |
| **25_at_T2-T1** | 2 | 0 | 0 | 1 | 1 | 4 | 2 | 198.5 | *ABL1* |
| **7027_at_T2-T1** | 10 | 2 | 58.9 | 0.910996 | 1 | 4 | 1 | 349 | *TFDP1* |
| **59338_at_T1** | 9 | 1 | 38.11111 | 0.998846 | 1 | 4 | 1 | 147 | *PLEKHA1* |
| **220042_at_T1** | 10 | 1 | 58.2 | 0.983503 | 1 | 4 | 0 | 0 | *DDIAS* |
| **8204_at_T2** | 10 | 4 | 128.3 | 0.922052 | 1 | 5 | 9 | 117 | *NRIP1* |
| **10645_at_T1** | 7 | 5 | 122.1429 | 0.72602 | 1 | 5 | 5 | 43.8 | *CAMKK2* |
| **4118_at_T2** | 10 | 5 | 127.3 | 0.881897 | 1 | 5 | 4 | 114.5 | *MAL* |
| **29094_at_T1** | 10 | 5 | 162.7 | 0.846459 | 1 | 5 | 2 | 67 | *LGALSL* |
| **5734_at_T1** | 5 | 2 | 62.2 | 0.97423 | 1 | 5 | 1 | 79 | *PTGER4* |
| **55752_at_T2-T1** | 4 | 2 | 89.25 | 0.905883 | 1 | 5 | 1 | 84 | *SEPTIN11* |
| **660_at_T1** | 10 | 3 | 109.9 | 0.931958 | 1 | 5 | 0 | 0 | *BMX* |
| **51202_at_T2** | 3 | 2 | 39.66667 | 0.753919 | 1 | 5 | 0 | 0 | *DDX47* |
| **8631_at_T2-T1** | 10 | 2 | 139.8 | 0.981072 | 1 | 5 | 0 | 0 | *SKAP1* |
| **3267_at_T1** | 10 | 2 | 108.8 | 0.938666 | 2 | 5.5 | 1 | 33 | *AGFG1* |
| **286333_at_T2-T1** | 10 | 5 | 87 | 0.7928 | 1 | 6 | 6 | 197.5 | *FAM225A* |
| **23569_at_T2** | 8 | 2 | 60.875 | 0.937367 | 1 | 6 | 6 | 155.1667 | *PADI4* |
| **90_at_T2-T1** | 2 | 1 | 80.5 | 0.979691 | 1 | 6 | 2 | 150.5 | *ACVR1* |
| **5028_at_T2-T1** | 7 | 1 | 128 | 0.999708 | 1 | 6 | 2 | 339 | *P2RY1* |
| **165_at_T2-T1** | 1 | 1 | 141 | 0.914608 | 1 | 6 | 1 | 159 | *AEBP1* |
| **391356_at_T2** | 1 | 0 | 0 | 1 | 1 | 6 | 1 | 325 | *PTRHD1* |
| **56888_at_T1** | 9 | 1 | 81.11111 | 0.991334 | 1 | 6 | 0 | 0 | *KCMF1* |
| **56670_at_T1** | 9 | 2 | 31.11111 | 0.860709 | 1 | 7 | 6 | 5.833333 | *SUCNR1* |
| **10810_at_T2** | 6 | 5 | 107.3333 | 0.819078 | 1 | 7 | 4 | 76.25 | *WASF3* |
| **116362_at_T1** | 10 | 4 | 48.1 | 0.808501 | 1 | 7 | 2 | 12.5 | *RBP7* |
| **5905_at_T1** | 2 | 1 | 39.5 | 0.799112 | 1 | 7 | 2 | 29 | *RANGAP1* |
| **51304_at_T1** | 9 | 3 | 135.3333 | 0.976526 | 1 | 7 | 0 | 0 | *ZDHHC3* |
| **283897_at_T2-T1** | 4 | 1 | 29.75 | 0.827281 | 1 | 7 | 0 | 0 | *C16orf54* |
| **490_at_T2** | 5 | 1 | 64.8 | 0.920852 | 1 | 7 | 0 | 0 | *ATP2B1* |
| **83541_at_T1** | 7 | 3 | 81.71429 | 0.898053 | 2 | 7.5 | 5 | 41.4 | *FAM110A* |
| **6498_at_T2-T1** | 8 | 3 | 137.625 | 0.918772 | 1 | 8 | 1 | 352 | *SKIL* |
| **128611_at_T2-T1** | 7 | 2 | 26.57143 | 0.892391 | 1 | 8 | 1 | 191 | *ZNF831* |
| **10150_at_T1** | 9 | 4 | 49.11111 | 0.835823 | 1 | 8 | 0 | 0 | *MBNL2* |
| **27334_at_T2-T1** | 10 | 4 | 147.5 | 0.95489 | 1 | 8 | 0 | 0 | *P2RY10* |
| **217_at_T1** | 10 | 3 | 169.1 | 0.953349 | 1 | 9 | 2 | 13 | *ALDH2* |
| **79041_at_T2** | 5 | 1 | 40.2 | 0.9636 | 1 | 9 | 1 | 272 | *TMEM38A* |
| **375759_at_T1** | 4 | 1 | 21.75 | 0.94688 | 1 | 10 | 4 | 61 | *C9orf50* |
| **55332_at_T2** | 8 | 2 | 133.75 | 0.953284 | 1 | 10 | 1 | 253 | *DRAM1* |
| **101927438_at_T2** | 2 | 0 | 0 | 1 | 1 | 10 | 0 | 0 | *LINC01800* |
| **129642_at_T2** | 10 | 3 | 137.4 | 0.980202 | 2 | 10.5 | 5 | 117.6 | *MBOAT2* |
| **2717_at_T2-T1** | 1 | 0 | 0 | 1 | 1 | 11 | 0 | 0 | *GLA* |
| **26509_at_T1** | 10 | 1 | 62.2 | 0.98613 | 1 | 13 | 1 | 36 | *MYOF* |
| **23344_at_T2-T1** | 10 | 3 | 50 | 0.907102 | 1 | 14 | 0 | 0 | *ESYT1* |
| **28503_at_T2-T1** | 9 | 7 | 74.11111 | 0.613662 | 0 | 0 | 9 | 200.4444 | *IGHD2-15* |
| **6897_at_T2** | 10 | 6 | 129.5 | 0.813632 | 0 | 0 | 9 | 87.77778 | *TARS1* |
| **8804_at_T2-T1** | 9 | 9 | 118.1111 | 0.568961 | 0 | 0 | 8 | 203.25 | *CREG1* |
| **129642_at_T2-T1** | 10 | 9 | 63.1 | 0.478666 | 0 | 0 | 7 | 193.5714 | *MBOAT2* |
| **22915_at_T2** | 10 | 6 | 30.1 | 0.635417 | 0 | 0 | 7 | 102.2857 | *MMRN1* |
| **116362_at_T2-T1** | 10 | 6 | 119.1 | 0.836741 | 0 | 0 | 7 | 200.5714 | *RBP7* |
| **3695_at_T2-T1** | 8 | 8 | 87.75 | 0.565565 | 0 | 0 | 6 | 179.3333 | *ITGB7* |
| **6897_at_T1** | 10 | 7 | 119.4 | 0.760882 | 0 | 0 | 6 | 13.83333 | *TARS1* |
| **28410_at_T1** | 10 | 7 | 126 | 0.799303 | 0 | 0 | 6 | 34.5 | *IGHV3-72* |
| **57561_at_T2** | 7 | 7 | 208 | 0.656849 | 0 | 0 | 6 | 132.3333 | *ARRDC3* |
| **3267_at_T2** | 10 | 6 | 52.7 | 0.670924 | 0 | 0 | 6 | 107.6667 | *AGFG1* |
| **440712_at_T2-T1** | 10 | 6 | 69.1 | 0.738946 | 0 | 0 | 6 | 178.3333 | *RHEX* |
| **116236_at_T2-T1** | 10 | 6 | 56.2 | 0.703683 | 0 | 0 | 6 | 216.3333 | *ABHD15* |
| **122618_at_T2** | 10 | 5 | 72 | 0.835392 | 0 | 0 | 9 | 100.2222 | *PLD4* |
| **329_at_T2-T1** | 10 | 5 | 180.5 | 0.924975 | 0 | 0 | 9 | 202 | *BIRC2* |
| **8460_at_T1** | 8 | 2 | 137.75 | 0.959781 | 0 | 0 | 8 | 43.375 | *TPST1* |
| **27032_at_T1** | 8 | 5 | 114.25 | 0.825349 | 0 | 0 | 7 | 25.57143 | *ATP2C1* |
| **29094_at_T2** | 10 | 5 | 47.6 | 0.72626 | 0 | 0 | 7 | 122.8571 | *LGALSL* |
| **643733_at_T2-T1** | 7 | 4 | 50 | 0.727492 | 0 | 0 | 7 | 178 | *CASP4LP* |
| **4481_at_T2-T1** | 10 | 3 | 179.4 | 0.978269 | 0 | 0 | 7 | 212.8571 | *MSR1* |
| **64805_at_T2** | 10 | 2 | 85.7 | 0.949125 | 0 | 0 | 7 | 102.1429 | *P2RY12* |
| **91_at_T2** | 9 | 5 | 196.2222 | 0.885988 | 0 | 0 | 6 | 121.1667 | *ACVR1B* |
| **4048_at_T2** | 7 | 5 | 149.7143 | 0.839172 | 0 | 0 | 6 | 123.6667 | *LTA4H* |
| **23064_at_T1** | 10 | 4 | 173.9 | 0.960072 | 0 | 0 | 6 | 36.83333 | *SETX* |
| **2624_at_T1** | 7 | 4 | 127.5714 | 0.829642 | 0 | 0 | 6 | 57 | *GATA2* |
| **4481_at_T2** | 10 | 4 | 97.9 | 0.858685 | 0 | 0 | 6 | 105.1667 | *MSR1* |
| **64746_at_T2** | 10 | 4 | 99.1 | 0.815552 | 0 | 0 | 6 | 105.6667 | *ACBD3* |
| **64168_at_T2** | 7 | 4 | 99.57143 | 0.852609 | 0 | 0 | 6 | 127.8333 | *NECAB1* |
| **9551_at_T1** | 8 | 3 | 38.25 | 0.784139 | 0 | 0 | 6 | 40.16667 | *ATP5MF* |
| **23569_at_T1** | 8 | 3 | 92.75 | 0.912001 | 0 | 0 | 6 | 52 | *PADI4* |
| **8754_at_T2** | 10 | 3 | 103.7 | 0.915786 | 0 | 0 | 6 | 92.33333 | *ADAM9* |
| **643733_at_T2** | 7 | 3 | 34.14286 | 0.770994 | 0 | 0 | 6 | 117.1667 | *CASP4LP* |
| **440712_at_T1** | 10 | 2 | 80.5 | 0.9715 | 0 | 0 | 6 | 8.666667 | *RHEX* |
| **56833_at_T2** | 10 | 2 | 130.9 | 0.974447 | 0 | 0 | 6 | 84 | *SLAMF8* |
| **6604_at_T2-T1** | 6 | 1 | 24 | 0.902891 | 0 | 0 | 6 | 218.1667 | *SMARCD3* |
| **388387_at_T2** | 10 | 7 | 150.5 | 0.843036 | 0 | 0 | 5 | 132.4 | *LINC00671* |
| **5627_at_T2** | 10 | 7 | 79.6 | 0.677623 | 0 | 0 | 5 | 148.6 | *PROS1* |
| **221037_at_T2-T1** | 10 | 7 | 70.5 | 0.656654 | 0 | 0 | 5 | 229.8 | *JMJD1C* |
| **3600_at_T2** | 10 | 6 | 64.4 | 0.683535 | 0 | 0 | 5 | 101.6 | *IL15* |
| **25777_at_T2-T1** | 8 | 6 | 109.5 | 0.691743 | 0 | 0 | 5 | 182.4 | *SUN2* |
| **4973_at_T2-T1** | 10 | 6 | 140.4 | 0.861604 | 0 | 0 | 5 | 192.2 | *OLR1* |
| **283989_at_T2-T1** | 9 | 7 | 69.33333 | 0.673472 | 0 | 0 | 4 | 231.5 | *TSEN54* |
| **283989_at_T1** | 9 | 6 | 88.22222 | 0.761278 | 0 | 0 | 4 | 33.75 | *TSEN54* |
| **3753_at_T2** | 10 | 6 | 92.7 | 0.810749 | 0 | 0 | 4 | 133 | *KCNE1* |
| **23446_at_T2-T1** | 10 | 6 | 192.2 | 0.905076 | 0 | 0 | 4 | 182.5 | *SLC44A1* |
| **79887_at_T2-T1** | 10 | 6 | 110.5 | 0.804308 | 0 | 0 | 4 | 268.75 | *PLBD1* |
| **116931_at_T1** | 9 | 6 | 50.88889 | 0.66105 | 0 | 0 | 3 | 61.66667 | *MED12L* |
| **28623_at_T2** | 10 | 6 | 60.9 | 0.711317 | 0 | 0 | 3 | 142.3333 | *TRBJ2-6* |
| **5110_at_T2** | 10 | 6 | 71.9 | 0.703327 | 0 | 0 | 3 | 185 | *PCMT1* |
| **27250_at_T2-T1** | 10 | 6 | 78.5 | 0.724368 | 0 | 0 | 3 | 230.6667 | *PDCD4* |
| **759_at_T2-T1** | 10 | 6 | 150.7 | 0.869733 | 0 | 0 | 3 | 244.3333 | *CA1* |
| **56833_at_T1** | 10 | 7 | 150.3 | 0.784106 | 0 | 0 | 2 | 21 | *SLAMF8* |
| **1668_at_T1** | 10 | 7 | 183.3 | 0.858087 | 0 | 0 | 2 | 23.5 | *DEFA3* |
| **7779_at_T2** | 10 | 7 | 184.7 | 0.831502 | 0 | 0 | 2 | 149 | *SLC30A1* |
| **80853_at_T2-T1** | 10 | 7 | 163.6 | 0.85546 | 0 | 0 | 2 | 200 | *KDM7A* |
| **65010_at_T1** | 10 | 6 | 72.8 | 0.767165 | 0 | 0 | 2 | 17 | *SLC26A6* |
| **3005_at_T2** | 10 | 6 | 213.8 | 0.921842 | 0 | 0 | 2 | 93 | *H1-0* |
| **93978_at_T2** | 10 | 6 | 158.7 | 0.905189 | 0 | 0 | 2 | 133 | *CLEC6A* |
| **5166_at_T2** | 9 | 6 | 188.1111 | 0.92959 | 0 | 0 | 2 | 136.5 | *PDK4* |
| **4680_at_T2-T1** | 10 | 6 | 97.8 | 0.830154 | 0 | 0 | 2 | 174 | *CEACAM6* |
| **167555_at_T2-T1** | 10 | 6 | 188.7 | 0.87448 | 0 | 0 | 2 | 312.5 | *FAM151B* |
| **56670_at_T2-T1** | 9 | 6 | 208.7778 | 0.897353 | 0 | 0 | 2 | 322 | *SUCNR1* |
| **286333_at_T2** | 10 | 8 | 105.8 | 0.695544 | 0 | 0 | 1 | 96 | *FAM225A* |
| **91746_at_T2** | 10 | 7 | 227.6 | 0.915621 | 0 | 0 | 1 | 257 | *YTHDC1* |
| **23328_at_T2-T1** | 10 | 7 | 153 | 0.777421 | 0 | 0 | 1 | 351 | *SASH1* |
| **80832_at_T1** | 10 | 6 | 259.5 | 0.947835 | 0 | 0 | 1 | 10 | *APOL4* |
| **51523_at_T1** | 10 | 6 | 135.3 | 0.784224 | 0 | 0 | 1 | 21 | *CXXC5* |
| **83938_at_T1** | 10 | 6 | 125.5 | 0.84863 | 0 | 0 | 1 | 23 | *LRMDA* |
| **566_at_T2-T1** | 10 | 6 | 149.5 | 0.883522 | 0 | 0 | 1 | 107 | *AZU1* |
| **1088_at_T2-T1** | 10 | 6 | 32.8 | 0.629051 | 0 | 0 | 1 | 108 | *CEACAM8* |
| **8724_at_T2-T1** | 10 | 6 | 93.4 | 0.773593 | 0 | 0 | 1 | 354 | *SNX3* |
| **3779_at_T2-T1** | 10 | 6 | 219.2 | 0.897535 | 0 | 0 | 1 | 363 | *KCNMB1* |
| **91746_at_T2-T1** | 10 | 6 | 325.3 | 0.991317 | 0 | 0 | 1 | 462 | *YTHDC1* |
| **128346_at_T2** | 10 | 8 | 82.5 | 0.65019 | 0 | 0 | 0 | 0 | *C1orf162* |
| **51056_at_T2** | 10 | 7 | 234.3 | 0.859577 | 0 | 0 | 0 | 0 | *LAP3* |
| **10079_at_T1** | 10 | 7 | 234.6 | 0.924114 | 0 | 0 | 0 | 0 | *ATP9A* |
| **3680_at_T2** | 10 | 7 | 255.9 | 0.924343 | 0 | 0 | 0 | 0 | *ITGA9* |
| **79674_at_T1** | 10 | 7 | 257 | 0.906088 | 0 | 0 | 0 | 0 | *VEPH1* |
| **80832_at_T2** | 10 | 7 | 262.3 | 0.957985 | 0 | 0 | 0 | 0 | *APOL4* |
| **7071_at_T2-T1** | 10 | 7 | 300.4 | 0.976585 | 0 | 0 | 0 | 0 | *KLF10* |
| **166_at_T1** | 9 | 6 | 60.88889 | 0.620548 | 0 | 0 | 0 | 0 | *TLE5* |
| **2681_at_T1** | 9 | 6 | 122.4444 | 0.853881 | 0 | 0 | 0 | 0 | *GGTA1* |
| **51477_at_T2** | 10 | 6 | 123.6 | 0.866065 | 0 | 0 | 0 | 0 | *ISYNA1* |
| **128372_at_T2-T1** | 10 | 6 | 158.8 | 0.803898 | 0 | 0 | 0 | 0 | *OR6N1* |
| **10970_at_T1** | 10 | 6 | 191.8 | 0.878235 | 0 | 0 | 0 | 0 | *CKAP4* |
| **967_at_T2** | 9 | 6 | 194 | 0.854897 | 0 | 0 | 0 | 0 | *CD63* |
| **974_at_T2** | 10 | 6 | 195.4 | 0.924924 | 0 | 0 | 0 | 0 | *CD79B* |
| **383_at_T1** | 10 | 6 | 195.5 | 0.889603 | 0 | 0 | 0 | 0 | *ARG1* |
| **7226_at_T2-T1** | 10 | 6 | 201.4 | 0.917255 | 0 | 0 | 0 | 0 | *TRPM2* |
| **6272_at_T1** | 10 | 6 | 211.3 | 0.925028 | 0 | 0 | 0 | 0 | *SORT1* |
| **4318_at_T1** | 10 | 6 | 214.4 | 0.936923 | 0 | 0 | 0 | 0 | *MMP9* |
| **90550_at_T2-T1** | 10 | 6 | 217 | 0.884786 | 0 | 0 | 0 | 0 | *MCU* |
| **825_at_T1** | 10 | 6 | 255.7 | 0.945024 | 0 | 0 | 0 | 0 | *CAPN3* |

\# times selected indicates the number of times a given feature was selected as informative by each feature selection method. Mean rank indicates the rank for each feature averaged across every iteration that particular feature was selected. VIP for random forest represents the variable importance measure for each feature.
